# Sex differences in the genetic regulation of the blood transcriptome response to glucocorticoid receptor activation

**DOI:** 10.1101/2020.10.19.20213983

**Authors:** Sarah R. Moore, Thorhildur Halldorsdottir, Jade Martins, Susanne Lucae, Bertram Müller-Myhsok, Nikola S. Müller, Charlotte Piechaczek, Lisa Feldmann, Franz Joseph Freisleder, Ellen Greimel, Gerd Schulte-Körne, Elisabeth B. Binder, Janine Arloth

## Abstract

Substantial sex differences have been reported in the physiological response to stress at multiple levels, including the release of the stress hormone, cortisol. Here, we explore the genomic variants in 93 females and 196 males regulating the initial transcriptional response to cortisol via glucocorticoid receptor (GR) activation. Gene expression levels in peripheral blood were obtained before and after GR-stimulation with the selective GR agonist dexamethasone to identify differential expression following GR-activation. Sex stratified analyses revealed that while the transcripts responsive to GR-stimulation were mostly overlapping between males and females, the quantitative trait loci (eQTLs) regulation differential transcription to GR-stimulation were distinct. Sex-stratified eQTL SNPs (eSNPs) were located in different functional genomic elements and sex-stratified transcripts were enriched within postmortem brain transcriptional profiles associated with Major Depressive Disorder (MDD) specifically in males and females in the cingulate cortex. Female eSNPs were enriched among SNPs linked to MDD in genome wide association studies. Finally, transcriptional sensitive genetic profile scores derived from sex-stratified eSNPS regulating differential transcription to GR-stimulation were predictive of depression status and depressive symptoms in a sex-concordant manner in a child and adolescent cohort (*n* = 584). These results suggest potential of eQTLs regulating differential transcription to GR-stimulation as biomarkers of sex-specific biological risk for stress-related psychiatric disorders.

## INTRODUCTION

Robust sex differences have been reported for stress-related psychiatric disorders, including mood and anxiety disorders, schizophrenia, and post-traumatic stress disorder (PTSD) (Abel et al., 2010; Diflorio & Jones, 2010; Ramikie & Ressler, 2018; Salk et al., 2017). Beyond prevalence rates, consistent sex differences are observed in the age of onset, symptomology, comorbidities and responses to medication (Abel et al., 2010; Boyd et al., 2015; Ramikie & Ressler, 2018; Salk et al., 2017). For instance, major depressive disorder (MDD) demonstrates higher prevalence rates in women than in men (Salk et al., 2017) and women exhibit heightened vulnerability to mood symptoms in association with stress-induced inflammatory processes (Bekhbat & Neigh, 2018). Despite the accumulating evidence for sex differences in stress-related pathogenesis of psychiatric conditions, the etiological mechanisms responsible for these differences are not well understood. Elucidating sex-related factors that moderate stress susceptibility is critical for targeted prevention and treatment strategies.

Evidence suggests that a dysregulation of the hypothalamic-pituitary-adrenal (HPA) axis contributes to vulnerability to stress (Bale & Epperson, 2015; Bekhbat & Neigh, 2018; Gold, 2015; Stephens et al., 2016). Exposure to stressful environments or threat leads to the activation of the HPA axis, with release of hypothalamic corticotropin-releasing hormone (CRH) that in turn stimulates release of adrenocorticotropin from the pituitary into the peripheral circulation. This leads to the release of glucocorticoids (GC) from the adrenal cortex. GCs bind to mineralo- and glucocorticoid receptors (GR), with the GR regulating biological adaptations to chronic stressors (Matthews, 1998; Owen & Matthews, 2003; Reul & De Kloet, 1985). The GR is highly expressed in most tissues both peripherally and centrally. Activation of GR by GCs causes the translocation of GR from the cytoplasm to the nucleus (de Kloet et al., 2005). There it binds to glucocorticoid response elements (GREs) and regulates gene expression. The resulting biological cascade has broad biological effects, initiating physiological changes in the body for adaptation to threat, and also providing negative feedback regulation to the brain for recovery (Sapolsky et al., 2000).

Sex differences in the stress response have been amply demonstrated at the physiological, hormonal, and neuroinflammatory levels (Bale & Epperson, 2015; Bekhbat & Neigh, 2018). In human studies, sex differences have been reported in both physiological and emotional responses to standardized stress tests, such as the Trier Social Stress Test (Childs et al., 2010; Kelly et al., 2008; Liu et al., 2017). Importantly, these stress response indices demonstrate abnormalities following exposure to childhood trauma (Tiwari & Gonzalez, 2018) and in stress-related psychiatric disorders (Zorn et al., 2017). Thus, a better understanding of sex differences in the stress response may inform the sex-biased pathways to stress- and trauma-related psychiatric disorders.

Sex differences in the stress response have largely been attributed to gonadal hormone changes. Sex chromosomes determine gonad development and gonadal hormones then alter regulatory pathways affecting the transcriptome and epigenome in sex-specific ways (Morrison et al., 2014). Indeed, the transcriptome (Aguet et al., 2020a, 2020b; Ellegren & Parsch, 2007; Jansen et al., 2014) and epigenome (Jessen & Auger, 2011; Sugathan & Waxman, 2013) are highly sex-specific. Animal models have shown that transcriptional changes due to stress exposure are sex-specific in the hippocampus (Rowson et al., 2019) and hypothalamus (Karisetty et al., 2017). Sex-specificity of the transcriptome extends to transcriptional signatures of MDD in humans (Brivio et al., 2020). For instance, MDD-associated transcriptional networks across brain regions are highly disparate between males and females, with sex-stratified results converging with sex differences in a mouse model of chronic social stress (Labonté et al., 2017). Taken together, these findings suggest a role for sex differences in genome function and regulation in sex-specific etiologies of stress-related disorders (Khramtsova et al., 2019).

Although allele frequencies do not differ between males and females across the autosomes (Boraska et al., 2012), GWAS sufficiently powered to allow stratification by sex have demonstrated the heterogeneity of genetic effects between males and females in association with complex traits (Khramtsova et al., 2019). Genetic variants may indeed show sex bias in their regulation of gene expression, supported by identified autosomal sex-biased *cis*-expression quantitative trait loci (**eQTLs**) in whole blood (Aguet et al., 2020a; Yao et al., 2014). Thus, in addition to regulation across the genome by gonadal hormones, there may also be sex-specific influences of genetic variants on downstream epigenetic and regulatory elements. Targeting these sex differences in genetic regulation of stress pathways, in particular, may elucidate sex-specific pathways of risk for psychiatric disorders.

Previously, we explored genetic variants that regulate the **GR-response**, defined as the immediate transcriptional response to glucocorticoids in humans, in our design via administration of dexamethasone, a selective agonist for GR (Arloth, Bogdan, et al., 2015). By quantifying gene expression in peripheral blood at baseline and three hours post dexamethasone administration, we reported eQTLs which modulate the transcriptome response to GR-activation in men. The eQTL SNPs (**eSNPs**) were shown to be enriched among genetic variants associated with schizophrenia as well as MDD and to predict amygdala reactivity to threat (Arloth et al., 2015) as well as neurovascular-coupling related features of the brain stress response (Elbau et al., 2018). The transcripts regulated by these variants form tight co-expression networks. Using an animal model of exposure to adversity across development (Santarelli et al., 2017), we observed that different combinations of early and adult environments (supportive vs. stressful) substantially affect co-expression structure of these networks in a highly brain region-specific manner (Zimmermann et al., 2019). However, this set of eQTLs and regulated transcripts was identified in a male only cohort.

Given the above described sex differences in the stress-response as well as in the prevalence and manifestation of psychiatric disorders, we conducted a sex-stratified analysis of genetic regulation of the transcriptional response to GR-activation in peripheral blood cells. We found that while transcripts regulated by GR-activation were largely overlapping in males and females, genetic variants moderating these GR-induced transcriptional changes (**GR-eQTLs**) were mainly identified in only females or males, suggesting that distinct genetic features moderate the transcriptional response to GR-activation in the two sexes. The transcripts regulated by GR-eQTLs (**etranscripts**) were enriched among sex-stratified transcriptional signatures of MDD in post-mortem brain tissue (Labonté et al., 2017). Sex-stratified GR-eQTLs were enriched in GWAS signals for MDD. Transcriptional sensitive genetic profile scores derived from sex-stratified GR-eQTLs also predicted depression and depressive symptoms in an adolescent cohort in a sex-specific manner. Our results underline the importance of sex-stratified analyses in stress-induced gene-regulation for a better understanding of stress-related psychiatric disorders.

## RESULTS

Whole blood samples from 289 individuals (93 females [48 patients with depression and 45 healthy controls] and 196 males [81 patients with depression and 115 controls]) recruited at the Max Planck Institute of Psychiatry (MPIP) were analyzed for gene expression levels at baseline and three hours post stimulation by the selective GR-agonist dexamethasone (see Arloth et al. 2015), see Table 1 for description. 11,994 transcripts entered the analysis. Additionally, all samples were genotyped, with a total of 3.9 Million SNPs available for analysis. All analyses were conducted only on autosomes to allow comparison between males and females and controlled for age, case-control status, BMI and cellular heterogeneity using surrogate variables (*n*=3, see **Supplementary Figure 1**). **Figure 1** displays an overview of the data analysis and results outlined below.

**Table 1:** Clinical characteristics. For continuous data the mean ± standard error and for categorical data the categories separated by dashes are given for females and males.

|  | MPIP cohort |  | LMU cohort |  | recMDD cohort |  |
| --- | --- | --- | --- | --- | --- | --- |
| Sex | males | females | males | females | males | females |
| N | 196 | 93 | 201 | 383 | 255 | 312 |
| Age | 42.65±13.7 | 42.95±14.6 | 14.5±2.3 | 15.5±1.8 | 46.53±13.9 | 47.0±13.8 |
| BMI | 25.4±3.3 | 23.9±5.1 | NA | NA | 24.7±3.1 | 24±4.5 |
| N controls/cases | 115/81 | 45/48 | 115/86 | 235/148 | 78/177 | 114/198 |

**Figure 1.**
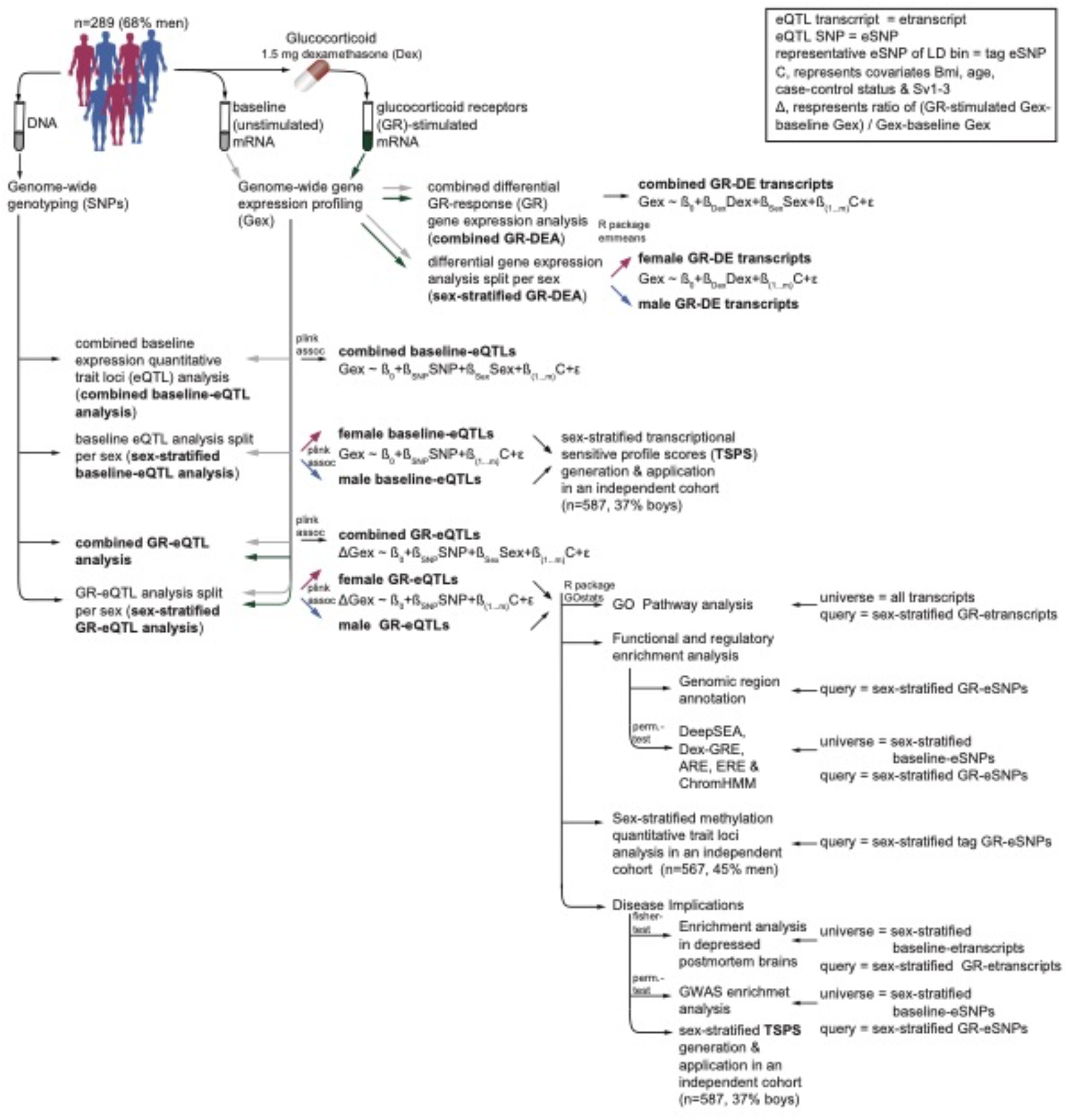
Flow of data collection and statistical analyses: Genome-wide genotyping and gene expression profiling was used to examine differential GR-response gene expression and expression quantitative trait loci in 1) combined and 2) sex-stratified analyses. Results were carried forward for functional interrogation and linkage to disease.

### GR-stimulated gene expression: comparison of males and females

First, we assessed the main effects of dexamethasone on gene transcription in a combined differential GR-response gene expression analysis (**combined GR-DEA**) in all participants controlling for sex (**Figure 1**). These results were then compared to differential gene expression analyses stratified by sex (**sex-stratified GR-DEA**), as well as a differential gene expression analysis testing the effect of sex on GR-stimulated changed in gene expression (see Supplementary Results and **Supplementary Table 1**). The combined GR-DEA identified 7,462 out of 11,994 autosomal transcripts to be significantly differentially expressed at an *FDR* of 0.05, and 2,352 transcripts (31.5%) to surpass an absolute log_2_ fold change (FC) threshold of 0.2 (see **Supplementary Table 2**). The majority of transcripts found to be regulated by dexamethasone in the combined GR-DEA were also identified in the sex-stratified GR-DEA, with few additional transcripts emerging (*n* = 253 in females and *n* = 15 in males; **Figure 2A**). Next, we assessed the consistency of the magnitude and direction of GR-DE changes across males and females (**Figure 2B-C, Supplementary Table 2**). Overall, larger log FCs were found in females (**Figure 2D** and **Supplementary Figure 2**). Further analyses supported that effects sizes, rather than direction, were moderated by sex, with consistent effect directions found in males and females (**Figure 2C-D**, see **Supplemental Results**). Sex-stratified GR-DEA effects were likely not driven by differences in dexamethasone serum levels. At the timepoint of the second blood draw, no differences in dexamethasone levels were observed between sexes in a subset of 162 males and 68 females (mean ln dexamethasone level = 2±0.25 in males and 1.92±0.93 in females, *p value* = 0.46). Thus, we conclude that sex differences in GR-response are largely due to magnitude of the transcriptome change rather than direction.

**Figure 2.**
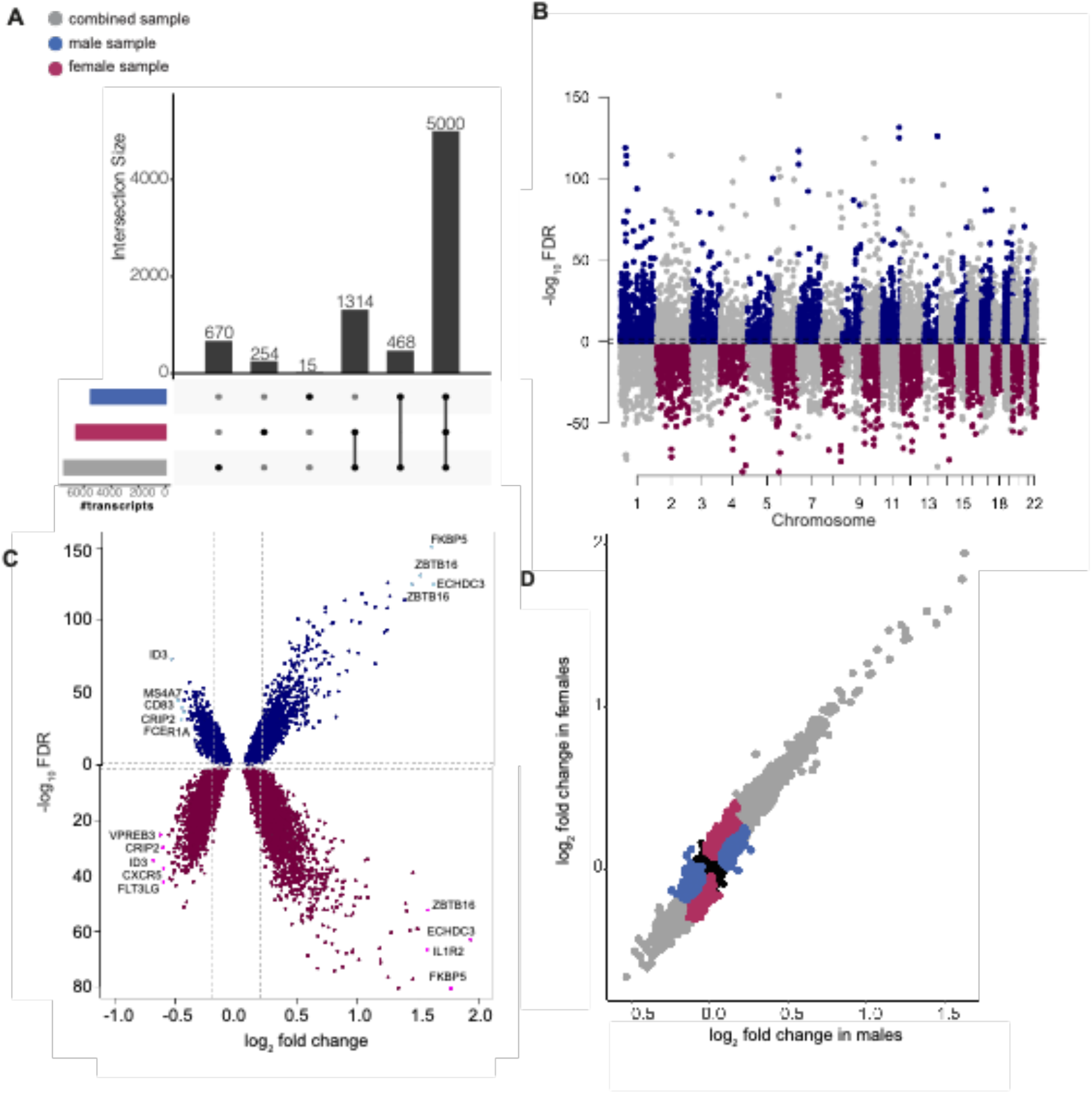
Differential GR-response gene expression analysis (GR-DEA): **A)** Upset plot displaying the overlapping significant transcripts identified in combined and sex-stratified GR-DEA. The majority of transcripts were found in both the combined and stratified analyses independently. **B)** Miami plot of results across 11,994 autosomal transcripts. Dashed lines indicate significance cut-off at an FDR of 5%. 6,568 GR-DE transcripts were significantly differentially regulated in females (*n* = 93 individuals; bottom panel) and 5,483 GR-DE transcripts in males (*n* = 196 individuals; top panel). **C)** Volcano plot of log_2_ fold change (x axis) by -log_10_*FDR*. Upper panel showing male GR-DE transcripts at an FDR of 0.05 with FCs ranging from 0.68 to 3.06. Lower panel showing female GR-DE transcripts with FCs ranging from 0.62 to 3.82. **D)** Scatterplot showing the difference in gene expression between post dexamethasone and baseline for males (y axis) and females (x axis) colored by identification in combined analysis (*n* = 5,000 transcripts), females (n=1568), males (*n* = 483), and neither females or males (*n* = 4,943). Significant results, whether supported in the combined analysis or limited to sex stratified analyses, are mainly limited to the upper right and lower left quadrants, supporting consistent effect directions between males and females.

### Sex differences in genetic regulation of GR-response

We next investigated sex differences in the genetic regulation of the transcriptional GR response. We focused on ***cis*-eQTLs**, which were defined as associations between SNPs and transcripts within a 1Mb window. C*is*-eQTL analyses were performed to identify **baseline eQTLs** (eSNPs significantly related to gene expression in unstimulated mRNA) and **GR-eQTLs** (eSNPs significantly related to the change in gene expression after GR stimulation). These analyses were carried out again in the combined sample (**combined baseline-eQTL analysis** and **combined GR-eQTL analysis**) and stratified by sex (**sex-stratified baseline-eQTL analysis** and **sex-stratified GR-eQTL analysis**). Although a cohort was not available to validate sex-stratified GR-eQTLs, we used publicly available data to validate sex-stratified baseline-eQTLs. We again focused on overlap of *cis* GR-eQTL effects in the sex-stratified analysis (i.e., common combinations of eSNPs and etranscripts), and the consistency of effect sizes and directions between males and females.

The combined GR-eQTL analysis identified 10,398 significant GR-eQTLs after multiple test correction, involving 717 etranscripts and 10,078 eSNPs. The 10,078 unique GR-eSNPs can be summarized into 747 uncorrelated GR-eSNP bins, i.e. sets of SNPs in linkage disequilibrium (LD) represented by a tag eSNP (see Methods and Arloth et al., 2015). These 747 tag GR-eSNP bins correspond to 804 GR-eQTL bins, i.e., eSNP bin-probe combinations, with some tag eSNPs associated with the expression of more than one transcript and are listed in **Supplementary Table 3**.

Next, the sex-stratified GR-eQTL analysis (**Figure 3A**) again indicated that effect directions were consistent between males and females (**Figure 3B**). In females, GR eQTLs were found for 648 eQTL bins comprising 613 etranscripts and 601 tag eSNP (**Supplementary Table 4)**. Slightly more eQTLs were identified in males with 705 eQTL bins involving 662 etranscripts and 668 tag eSNPs (**Supplementary Table 5**).

**Figure 3:**
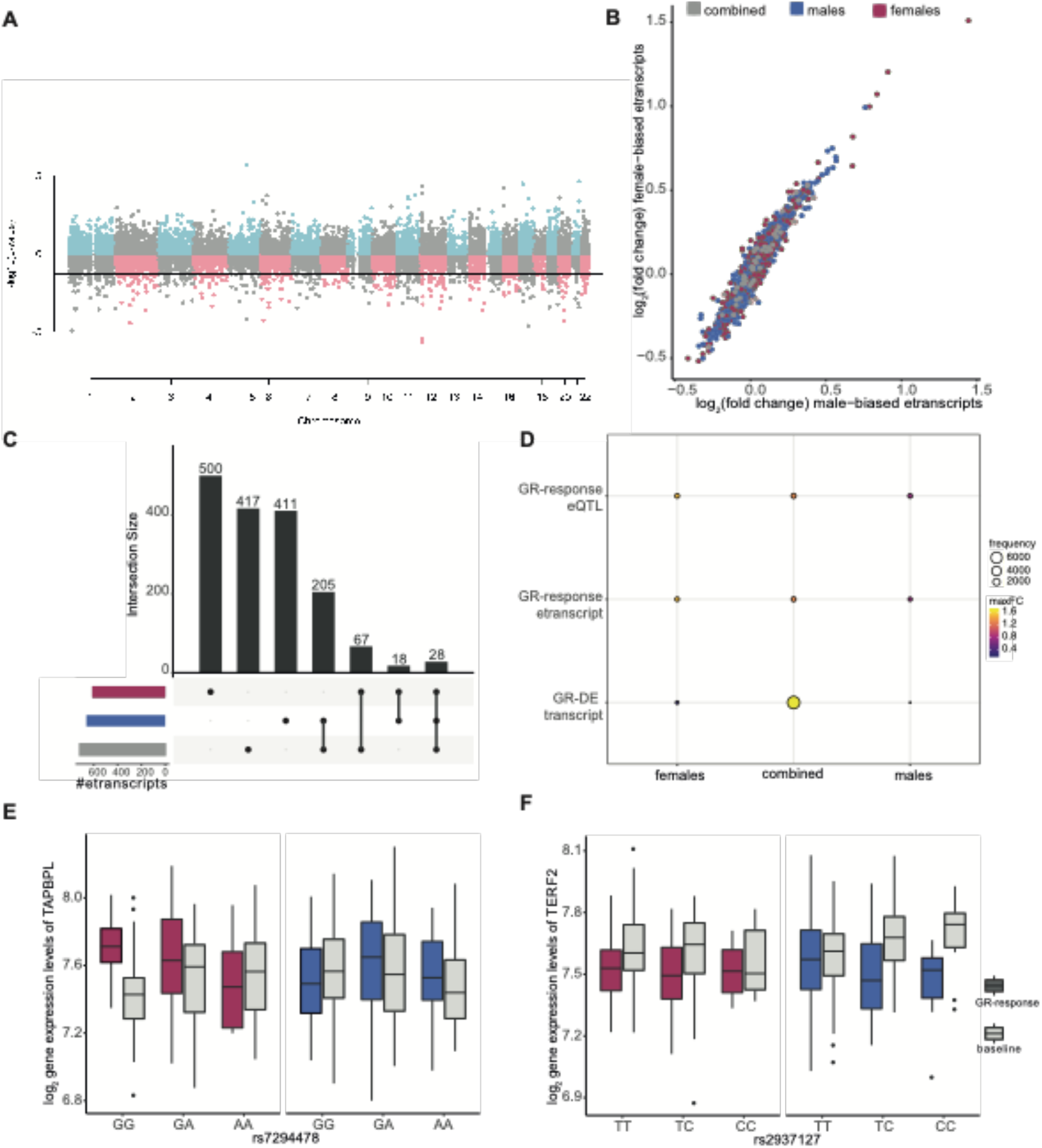
GR-response *cis*-eQTL analyses: **A)** Miami plot of eQTL results. Only the best eQTL per etranscript is plotted. Dashed lines indicate significance cut-off at an FDR of 5%.**B**) Mean log2 fold changes between post dexamethasone and baseline colored by identification of etranscripts in combined analysis (*n* = 46 transcripts), females (*n =* 567) or males (*n =* 616). The effects of the etranscripts for male and females were similar (Wilcoxon *p-value* =0.7). **C**) Upset plot displaying the overlapping significant GR-response etranscripts identified in combined analysis, males, and females. The majority of these transcripts were specific to females (91%, *n* = 193), whereas 68 (59%) transcripts were specific to males and 74 (57%) transcripts were found in the combined eQTL analysis. **D)** Balloon plot showing the frequency of transcripts found in 1) females but not the combined analysis, 2) the combined analysis, and 3) males but not the combined analysis, across GR-DE transcripts, etranscripts, and etranscript-eSNP pairs. In the GR-DE analysis, the majority of transcripts are identified in the combined analysis, whereas etranscripts and eSNP pairs (eQTLs) show more of an even distribution across females, combined, and males. Maximum fold changes were higher in female etranscripts relative to males. **E-F)** Boxplots of overlapping significant GR-DE transcripts and etranscripts. Gene expression is stratified by eSNP and shown for females and males. **E)** Tag eSNP rs7294478 is located in an intron of *C1RL-AS1* on chromosome 12. However, the eQTL effect was observed only in females on *TAPBPL* expression, which is located over 700 kb downstream. *TAPBPL* itself is one of the significant DR-DE genes identified in females (*FDR* = 0.00068 vs. *FDR* = 1 in males). **F)** The intronic tag eSNP rs2937127 demonstrates no effect in females, while in males the minor allele was associated with a down regulation of *TERF2* gene expression (*FDR* = 0.04). *TERF2* is located approximately 470 kb upstream of the tag eSNP, which is positioned in the gene *WWP2*.

By overlapping the female and male stratified results with the combined GR-eQTL analysis, we show that 34% of the male GR-response etranscripts (*n* = 233) and 16% of the female GR-response etranscripts (*n* = 95) were identified as etranscripts by the combined model (**Figure 3C**). Thus, in contrast to the GR-DEA results, the sets of etranscripts are largely non-overlapping (**Figure 3D** and **Supplementary Figure 3)**.

An example of a female GR-eQTL compared to males and to the combined sample is displayed in **Figures 3E-F**. Approximately 50% of etranscripts identified in the sex-stratified GR-eQTL analysis were also identified as sex-stratified GR-DEA transcripts (**Figure 3D**), with female etranscripts exhibiting larger log2FCs relative to males (see **Supplemental Results)**. We next compared enrichment of biological functions for GR etranscripts between males and females. Female etranscripts were enriched for regulation of natural killer cell mediated immunity and male etranscripts were enriched for regulation of cyclin-dependent protein kinase activity, positive regulation of extrinsic apoptotic signaling pathway, peptide metabolic processes, and other functions (see **Supplementary Table 7**). Additionally, we were able to validate the majority (86% male baseline eQTLs and 84% female baseline eQTLs) of baseline eQTLs in publicly available data (see **Supplementary Results**).

### Functional and regulatory context of sex-stratified GR-eSNPs

We next characterized the identified GR-eSNPs (unpruned) in terms of genomic location, regulatory features, and enrichment for sex hormone response elements. GR-eSNPs for females were significantly more likely to be located in distal intergenic regions (40.9%) compared to male GR-eSNPs (34.4%), see **Figure 4A (**fisher exact *p-value* = 1.4×10^−14^). GR-male eSNPs were significantly more likely to cluster in intronic regions (50% vs. 42.9% in first or other introns for GR male and GR-female eSNPs, respectively (fisher exact *p-value* = 3.4×10^−16^).

**Figure 4:**
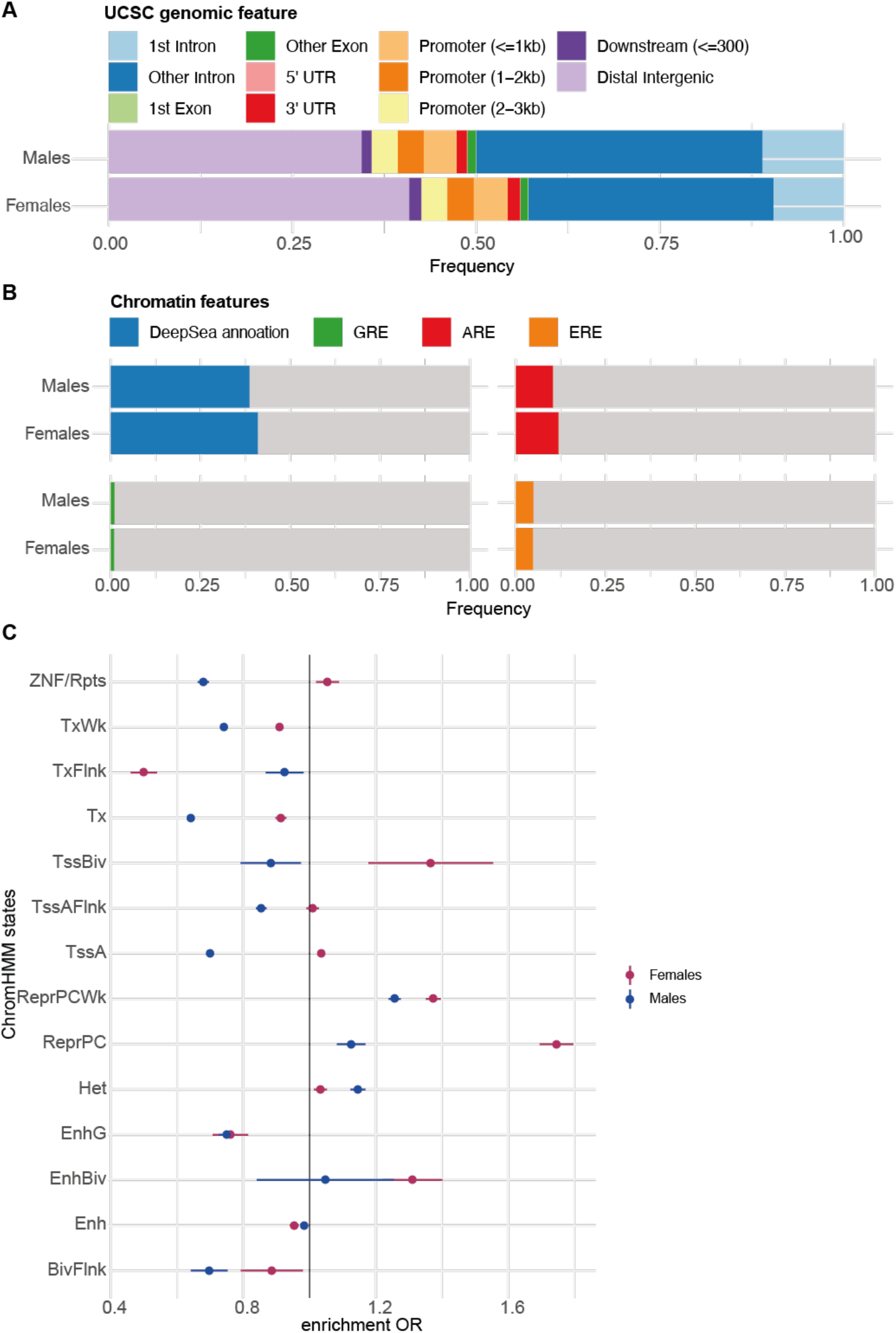
Sex stratified GR-response *cis-*eQTLs and chromatin annotation: **A**) Annotation of the genomic regions in which eSNPs are located. **B**) Bar plots showing the overlap of GR-eSNPs and DeepSea annotations, Remap transcript factors (AR and ER) and Encode GR-Chip peaks. **C**) Enrichment results for GR-response tag eSNPs and predicted ChromHMM states for sex-stratified tag eSNPs.

As eQTLs have previously been associated with regulatory regions (Fadason et al., 2018), we quantified enrichments of male and female GR-eSNP sets for regulatory features. For all tests, enrichment for sex-stratified GR-eSNPs were tested for significant enrichment compared to sex-stratified baseline eSNPs set as background to ensure sex-stratified effects were specific to eSNP regulation of the GR-response, specifically. Thus, male and female GR-eSNPs had to show significant enrichment relative to male and female baseline eSNPs. First, we used DeepSEA, a deep neural network pretrained with DNase-seq and ChIP-seq data from the ENCODE project, to predict the likelihood that GR-sex eSNPs exert regulatory effects on chromatin features. We found 8.4% of the GR female eSNPs (*n* = 500) with significantly overlapping DeepSEA features (*e-value* < 0.01) and 10.7% of the male GR eSNPs (*n* = 851), contained DeepSEA features (**Figure 4B**). Additionally, using GRE ChIP-Seq peaks from ENCODE lymphoblastoid cell lines treated with dexamethasone, we observed significant overlap within GR-binding sites (GREs) for female eSNPs (*n* = 58 out of 5586 eSNPs, enrichment *p-value* = 0.022, *OR* = 1.46, **Figure 4B**), but not male eSNPs.

To determine if the sex-stratified GR eSNPs are more likely to be located within sex hormone responsive regulatory elements, we calculated the number of eSNPs that are located within androgen response elements (AREs) and estrogen response elements (EREs), using data from Remap (see Methods). Of all 5,586 GR-female eSNPs, 4.89% (*n* = 273, **Figure 4B**) were located within EREs and 11.94 % (*n* = 667, **Figure 4B**) in AREs. For the 7,771 GR-male eSNPs, 4.95% (*n* = 382, **Figure 4B**) and 10.38% (*n* = 807, **Figure 4B**) were located within EREs and AREs, respectively. Enrichments for AREs and EREs were not statistically significant above sex-stratified baseline eQTLs. These results suggest that sex-stratified eSNPs may potentially be independent of the direct influence of sex hormones, in accordance with previous results (Khramtsova et al., 2019; Mayne et al., 2016).

Sex-stratified GR eSNPS were additionally tested for enrichment for hormone-related transcription factors (TFs) including ESR1, AR, and NR3C1 using Remap. Although both male and female sex-stratified GR-eSNPs and sex-stratified baseline eSNPs demonstrated significant enrichments across these TFs, the sex-stratified GR-eSNPs were not significantly enriched relative to sex-stratified baseline eSNPs. Testing the full remap database, we found significant enrichment of EZH2 and NR5A2 for female GR-eSNPs above baseline eSNPs, and significant enrichment of SND1 and EZH2 for male GR-eSNPs above baseline eSNPs.

Using the 15-state ChromHMM annotation of the Roadmap Epigenomics project (Chadwick, 2012), we observed that both female and male GR-eSNPs were enriched within repressed polycomb and bivalent enhancers across the tissue group of blood and T-cells (*n* = 14 cell lines), see **Figure 4C**. Female GR-eSNPs were enriched in ZNF genes and repeats, bivalent and poised transcription start sites (Tss), and active Tss (TssA and TssAFlnk), while male GR-eSNPs were depleted in Tss (**Figure 4C**). For the individual blood cell lines and enrichment *p-values*, see **Supplementary Figure 4**. All results were consistent whether using all eSNPs, or limiting the analysis to tag eSNPs, suggesting that results were not dependent on the structure of eSNPs in LD.

### Epigenetic modifications of sex-stratified GR eSNPs

As regulatory effects of sex-stratified GR-eSNPs may also act at the level of the epigenome, we explored links between sex-stratified GR-eSNPs and DNA methylation levels at baseline in an independent sample (recMDD cohort, see Methods) of 312 females and 255 males. We first performed sex-stratified methylation QTL (**meQTL**) analyses and identified 10,832,433 meQTLs in males comprising 163,238 CpGs and 2,94 million SNPs. Additionally, we found 12,691,324 meQTLs in females comprising 162,773 CpGs and 3,16 Mio SNPs at an FDR of 5% with 51.1% CpGs (*n* = 83,228) and 74.2% meQTL SNPs (meSNPs; *n* = 2.343.464) in common with the CpG identified in males only. Next, we quantified the number of sex-stratified GR-eSNP that are also significant meSNPs. Approximately half of both the female and male tag GR-eSNPs were meSNPs, i.e., 317 out of 601 female tag GR-eSNPs and 319 out of 668 male GR tag eSNPs (**Supplementary Figures 5A-C**). Thus, half of the sex-stratified eSNPs had additional associations with DNAm patterns.

**Figure 5:**
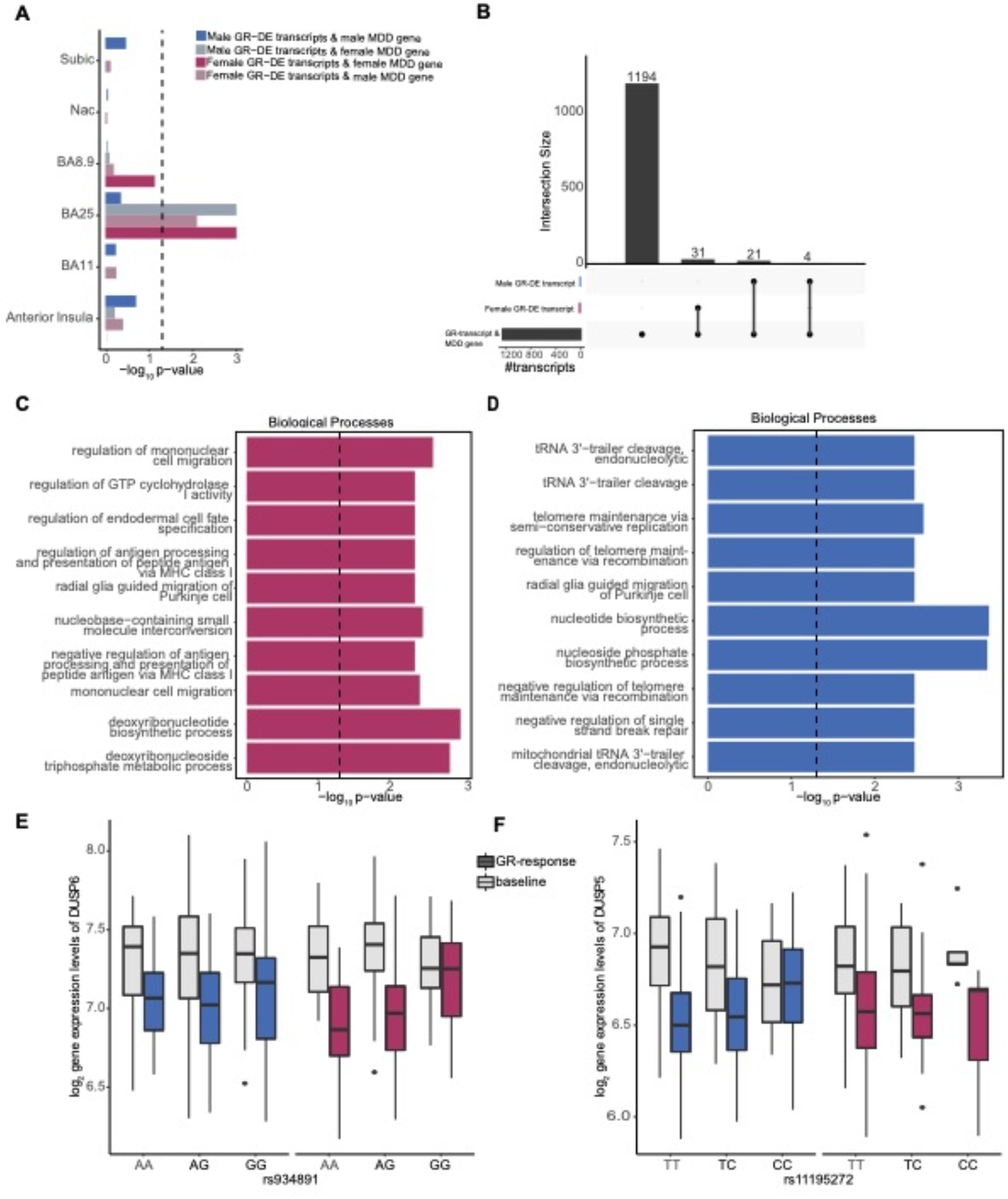
Sex-stratified GR-response etranscripts represented in MDD transcriptional patterns: **A**) Bar plot showing the significance of GR-etranscripts for enrichment in MDD transcriptional profiles in six brain regions. Both male and female GR-etranscripts were tested against male and female MDD transcriptional profiles. The black line indicates significance cut-off at a *p-value* of 0.05. **B)** Upset plot displaying the overlapping significant sex-stratified GR-response etranscripts with BA25 MDD-related transcripts. **C)** GO enrichment results for female etranscripts overlapping with BA25 MDD-related transcripts. **D)** GO enrichment results for male etranscripts overlapping with BA25 MDD-related transcripts. **E)** *DUSP6* example showing gene expression at baseline and post dexamethasone across genotypes of tag eSNP rs934891 for males and females (female *FDR* = 0.049). **F)** *DUSP5* example showing gene expression at baseline and post dexamethasone across genotypes of tag eSNP rs11195272 for males (male *FDR* = 0.046) and females.

### Disease Implications: sex-stratified GR-eQTLs predict depression and depressive symptoms

The potential disease relevance of the sex-stratified GR eQTLs was explored at three levels: enrichment in depression-related DE in human postmortem brain tissue, enrichment in GWAS associations for psychiatric disorders and traits and association of genetic profile scores weighted by sex-specific etranscript regulation.

#### Postmortem gene expression in major depression

We next explored whether sex-stratified GR-etranscripts and eSNPs were represented within previous findings on genetic risks and underpinnings of psychiatric disorders. First, sex-stratified GR-etranscripts (relative to sex-stratified baseline etranscripts) from blood were mapped to sex-stratified transcriptional differences in the brain in association with MDD (Labonté et al., 2017). GR-male etranscripts were significantly enriched (*FDR* < 5%) in Brodmann area (BA) 25 in female MDD genes, and GR-female etranscripts were enriched in BA25 in both male and female MDD genes, a critical area for mood disorders, targeted by deep brain stimulation in the treatment of depression (Bezchlibnyk et al., 2018), see **Figure 5A**. Neither male or female etranscripts were significantly enriched in other brain regions.

Sex-stratified etranscripts overlapping with female MDD-related BA25 transcripts included 37 female etranscripts and 27 male etranscripts (with 4 common etranscripts between males and females, **Figure 5B**). We tested whether these sex-stratified etranscripts exhibited functional pathway differences between males and females. Female overlapping etranscripts were significantly enriched for deoxyribonucleotide biosynthetic process and deoxyribonucleotide triphosphate metabolic process. Male overlapping etranscripts were enriched for nucleotide biosynthetic process (*OR* = 9.39, *p-value* = 0.0004) and nucleoside phosphate biosynthetic process (*OR* = 9.29, nominal *p-value* = 0.0004) (**Figure 5C-D**). Interestingly, Dual Specificity Protein Phosphatase 6 (*DUSP6)* was represented among female etranscripts, and *DUSP5* within male etranscripts, both members of an enzyme subfamily of dual-specificity MAP kinase phosphatases which are conserved in domain structure. *DUSP6*, in particular, was identified as a driving hub in MDD-related transcriptional networks (Labonté et al., 2017) and is involved in brain-related functions via inactivation of ERK pathways. Labonté and colleagues found that *DUSP6* was downregulated in female MDD subjects in BA25, and this pattern of downregulation was further supported by a mouse model of MDD in chronically stressed female mice. Although we found transcriptional effects in *DUSP6* to be common in males and females in response to GR activation, the eSNP effects were specific to females (**Figure 5E**), highlighting a sex specific mechanism regulating a common, downstream physiological pattern. *DUSP5*, similarly involved in ERK signaling in the brain, was also downregulated by GR activation in males and females in our GR-DE analysis, but with a specific eSNP effect for males (**Figure 5F**).

#### GWAS for psychiatric disorders and traits

To extend these results, we tested whether sex-stratified GR eSNPs were overrepresented among GWAS SNPs associated with psychiatric disorders using large-scale GWAS results of the Psychiatric Genomics Consortium (PGC), relative to sex-stratified baseline eSNPs. All enrichments were independent of LD as we used the top-associated SNP of the clumping procedure (i.e., the tag SNP). We detected a significant enrichment of female GR eSNPs (*n* = 598 tag eSNPs) compared to female baseline eSNPs (*n* = 1,074 tag eSNP) with SNPs at a nominal GWAS *p-value* cutoff associated with MDD (fold enrichments = 1.15-1.88, permutation-based *FDRs* < 0.05, educational attainment (fold enrichment = 1.18, permutation-based *FDR* = 0.003), autism spectrum disorder (fold enrichment = 1.38, permutation-based FDR <0.001), attention-deficit/hyperactivity disorder (fold enrichment = 1.28, permutation-based *FDR* = 0.013), cannabis intake (fold enrichment = 1.26, permutation-based *FDR* = 0.012) and the cross-disorder analysis 2013 (fold enrichment = 1.5, permutation-based *FDR* = 0.007), see **Figure 6A**. For male GR eSNPs we did not identify an enrichment over male baseline eSNPs. In summary, female eSNPs regulating the GR response, but not male eSNPS, were significantly enriched in SNPs identified in relation to psychiatric disorders in large-scale GWAS studies.

**Figure 6.**
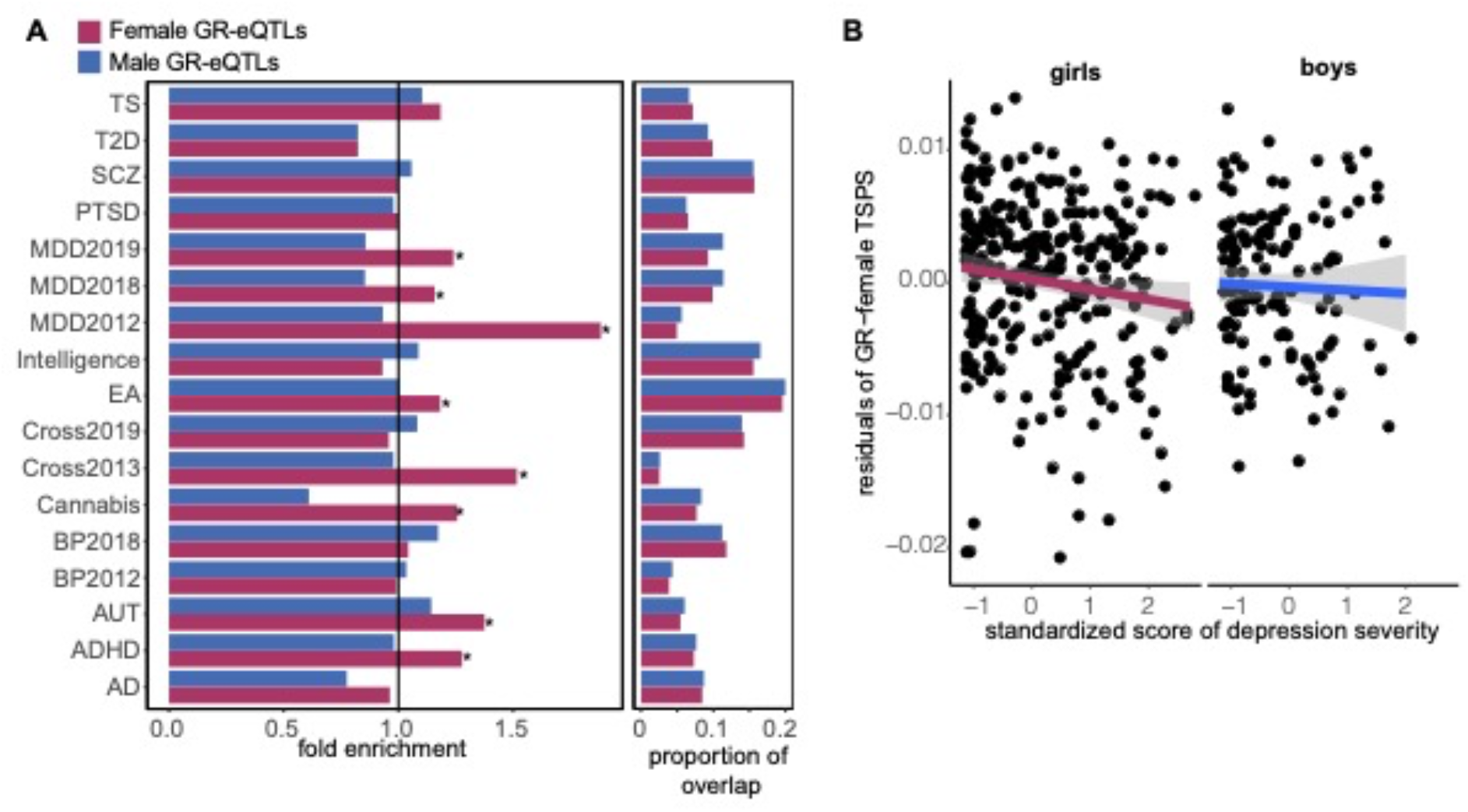
Sex-stratified GR-response eSNP associations with psychiatric disorders. **A)** Bar plot of enrichment results for GR-response tag eSNPs and GWAS SNPs. The black indicates a fold enrichment at 1 and a star indicates a permutation-based FDR < 0.05. AD = Alzheimer’s disease, ADHD=attention-deficit/hyperactivity disorder, AUT=autism spectrum disorders, BP = bipolar disorder, Cross = cross disorder analysis, EA=educational attainment, MDD= major depressive disorder, PTSD= post-traumatic stress disorder, SCZ = schizophrenia, T2D= diabetes type 2, TS= Tourette syndrome. **B**) Association between residualized female TSPSs and standardized scores of severity of depressive symptoms computed in LMU cohort (girls: β = -7.98×10^−4^, SE = 4.04×10^−4^, *p-value* = 0.0496; boys: β = 1.1210^−3^, SE = 8.3×10^−4^, *p-value* = 0.18)

#### Sex-specific genetic profile scores

Given the highly distinct sets of genetic variants regulating the GR-response in males and females, we assessed whether the genetic variants of GR-sex eQTLs would be cumulatively associated with sex-stratified sensitivity for psychiatric disorders.

Transcriptional sensitive profile scores (TSPS) were calculated by summation of the GR-eQTL effects. The ‘sensitive’ allele is defined as the allele with the highest absolute eQTL effect, regardless of effect direction, such that a higher TSPS represents elevated sensitivity for a GR-moderated transcriptional response. We tested whether TSPS based on sex-stratified eSNPs (**sex-stratified TSPS**) were associated with depression and depressive symptoms. We applied sex-stratified TSPSs to a clinical cohort comprising 350 Caucasian children and adolescents 7–18 years old with a current diagnosis or history of MDD (67% girls) and 234 healthy control subjects (ages 7–18 years old) with no history of a psychiatric disorders (63% girls, see Methods). Female TSPS significantly predicted case control status for depression in girls (*p-value =* 0.0256, see **Figure 6B**), explaining 2.3% of the variance in depression. Both the male and female TSPS significantly predicted specific depressive symptoms in the respective sex (*p-values*< 0.05, see **Supplementary Figure 6)**. The specific depressive symptoms related to TSPS were different for males and females. For instance, female TSPS significantly related to irritability, loss of satisfaction, agitation, crying, suicidal ideation, feelings of failure, and self−dislike, whereas male TSPS significantly related to changes in appetite, self-deprecation, anhedonia, and loss of interest. Both TSPSs significantly relate to worthlessness. Overall, female depressive symptoms were more self-directed or brooding than male symptoms.

Taken together, we found connections between sex-stratified eSNPs regulating the GR response and 1) transcriptional patterns in the brain in relation to MDD in women, 2) SNPs associated with psychiatric disorders, and 3) depression status and symptoms in a developmental cohort. Female eSNPs, in particular, were enriched in SNPs identified in psychiatric disorders, and as a cumulative score, were predictive of case-control status. Thus, sex-stratified eSNPs regulating the GR response may have relevance for the etiology of psychiatric disorders and implicate biological risk for their development in response to stress exposure.

## DISCUSSION

Sexual dimorphism in the stress response is well established, but how these sex differences are genetically regulated and linked to sex-specific risk for psychiatric conditions are unknown. Here, we explored potential sex differences in regulation of the stress response by comparing GC induced changes in gene transcription and *cis* genetic regulation of these changes in males and females. We find that sex differences in the transcriptomic GR-response are largely due to females demonstrating stronger effects of GR activation in terms of up and down regulation of transcripts, rather than differences in the direction of effects. However, the genetic regulation of the transcriptomic GR-response was highly disparate between sexes, with males and females demonstrating distinct sets of genetic variants corresponding to distinct patterns of regulatory features. Genes differentially expressed to GR activation in blood in males and females also demonstrated sex-specific patterns in postmortem brain of female patients with depression, and female GR-eQTLs were enriched among SNPs identified in large scale GWAS studies in relation to psychiatric disorders. Moreover, sex-stratified TSPSs created from GR sex-biased eSNPs predicted depression status and depressive symptoms in a clinical cohort of children and adolescents. Taken together, these findings have implications for identifying genetic sensitivity factors for males and females, corresponding to sex-specific biological susceptibility to stress exposure and stress-related psychiatric disorders.

Male and female GR-eQTLs could emerge due to direct genetic effects within the binding sites of GR, as well as due to epigenetic mechanisms at the level of chromatin (Lindén et al., 2017). To explore epigenetic mechanisms in relation to sex-stratified GR-eQTLs, we performed an integrative analysis of epigenetic states, including overlap of eSNPs with GR and sex hormone binding sites and linkage to sex-stratified SNP effects on DNAm (meQTLs). For both female and male GR-eQTLs, we found enrichment for regulatory chromatin features, but with sex-specific enrichments. Male and female GR-eQTLs which overlapped with sex hormone response elements were not enriched above male and female baseline eQTLs, suggesting that male and female GR genetic regulation may be independent of direct influences of sex hormones. We found that a substantial proportion (about half) of etranscripts regulated by sex-stratified GR-eQTLs were linked to sex-stratified meQTLs. Together, these results suggest that male and female GR-eQTLs have distinct downstream regulatory effects upon GR pathways and are also associated with sex differences in DNA methylation status, which may be important for sex differences in gene expression. Further, these regulatory effects appear to be, at least in part, independent of circulating sex hormones.

Previously, the study of biological differences between males and females largely targeted brain organization and regulation by sex hormones. More recently, attention is being paid to growing evidence in favor of genetic and epigenetic regulation of sexual dimorphism in behavior (Ratnu et al., 2017). By activating GR to directly assess genetic regulation of the stress response in males and females separately, our results add to a growing body of literature highlighting sex differences in gene expression and genetic regulation (Dimas et al., 2012; Gershoni & Pietrokovski, 2017; Jansen et al., 2014; Labonté et al., 2017; Mayne et al., 2016). In contrast to much of the work on the genomics of sex differences, we find that males and females differ in genetic regulation outside of the X and Y chromosomes. Thus, this work suggests that the genetic regulation of sex differences in stress responding extends tp the autosomes, and highlights the need for further work to understand the sex-specific genetic and epigenetic architecture underlying susceptibility to stress-related disorders.

Male and female GR-DE transcripts and etranscripts that were regulated by GR-eQTLs were found to be enriched for genes previously reported as sex-specific MDD transcriptional signatures in the brain. For these sets of significantly enriched genes identified in blood, their representation in the brain was not specific to males or females, despite the fact that these neural transcriptional signatures showed strong sex specificity in postmortem brain (Labonté et al., 2017). These results echo additional results presented in Labonte et al., namely, that although the transcriptional correlates of MDD in the brain were highly disparate between males and females, the downstream pathways of stress susceptibility converged. Interestingly, the enrichments were restricted to DE transcripts in BA25, the subgenual cingulate region, a brain area implicated in the pathophysiology of major depression and a target for deep brain stimulation as treatment for therapy resistant forms of this disease (Mayberg et al., 2005).

We have previously shown that GR-eQTLs in males are enriched among genetic variants associated with risk for psychiatric disorders, including MDD and SCZ (Arloth et al., 2015). The female GR-eQTLs we identified here were enriched for SNPs associated with MDD, EA, cannabis use, AUT, ADHD in large scale GWAS as well as cross disorder psychiatric risk (Cross-Disorder Group of the PGC et al., 2019; Ripke et al., 2013; Wray et al., 2018a). The selective enrichment of female GR-eQTLs in GWAS is interesting, as not all of the above disorders have a higher prevalence in girls or women. This would suggest, as also highlighted above, that sex-stratified GR-eQTLs target common pathways of risk, and emergence of disease is driven by a number of additional factors. A limitation of our enrichment analyses is that current GWAS mainly combine data from both sexes, even though a previous *post hoc* analysis of existing GWAS studies identified numerous significant loci that were driven by one sex or the other (Gilks et al., 2014) and another study identified genetic variants associated with MDD status in females only (Kang et al., 2020). Our results and these studies highlight that large-scale studies aimed at genetic discovery may benefit from modeling males and females separately.

Large scale GWAS have been used to derive polygenic risk scores, weighted by association strength to predict disease risk or better understand correlated biological features. However, these scores are limited by the fact that the underlying GWAS rely on heterogenous samples and imprecise measurement of complex phenotypes (Moore, 2017). Here, by manipulating the biological system of interest, we were able to preselect SNPs based on functional regulation. We weighted these SNPs by expression changes to dexamethasone, a direct gauge of the biological stress response shaped by an individual’s history of stress exposure, thus capturing genetic variability relevant to biological stress responding shaped across time, and regardless of environmental histories and idiosyncratic experiences.

These genetic sensitivity scores indeed demonstrated relevance to stress-related disorders. TSPS scores predicted depression status as well as symptoms in a sex-specific manner. Thus, both the genetic etiology, and the relations of these genetic sensitivity scores to MDD symptoms are specific to sex. Across sex-biased symptomology, higher scores of GR-eQTL dosage associated with larger biological responses to GR activation were associated with lower levels of depressive symptoms and status. This is in line with data from stress- and trauma research, showing that a blunted cortisol response in associated with higher risk for subsequent psychiatric disorders.

It is important to acknowledge a number of limitations to this study. First, our sample size, although considerably expanded relative to our previous report (Arloth et al., 2015), is still small for detecting small differences between males and females in the genetic regulation of GR-response gene expression and was imbalanced between males and females. Although GR activation by dexamethasone offers a substantial biological effect at the level of the transcriptome, replication of our results in an independent cohort is necessary. However, bootstrapping analysis indicated overall robustness of our finding (see **Results**). In addition, the majority of the sex-stratified baseline-eQTLs were also significant in public data, and thus we were well powered enough to replicate previous eQTL findings. Second, we were unable to control for timing of the menstrual cycle, and the use of birth control in women. Although this should be addressed in a replication, accounting for surrogate variables reflecting cell type proportions in our data should ameliorate any effects of this unwanted biological variation.

To the best of our knowledge, we are the first to report sex-stratified effects of GR activation in terms of differential gene expression in human blood. Moreover, this is the first study to identify male and female specific GR-eQTLs. In contrast to previous studies of biological sex differences in humans that often focus on sex chromosomes, we find significant and robust sex differences in terms of autosomal genetic variants in their regulation of the stress response with relevance to stress-related diseases. We report that these sex differences, both at the level of differential expression and genetic regulation of the GR-response, are large and robust enough that they emerge even in combined-sex models that control for sex. These findings highlight the need for careful examination of sex differences in the study of genetic risks and biological substrates of stress-related disorders.

## Data Availability

Data from MPIP gene expression experiment are deposited at the GEO repository under GEO:
GSE64930 and recMDD methylation under: GSE125105. Data analysis code is available at https://github.com/jArloth/sex-specific-GR-response-Analyses

## Supplementary Figures

**Figure S1:**
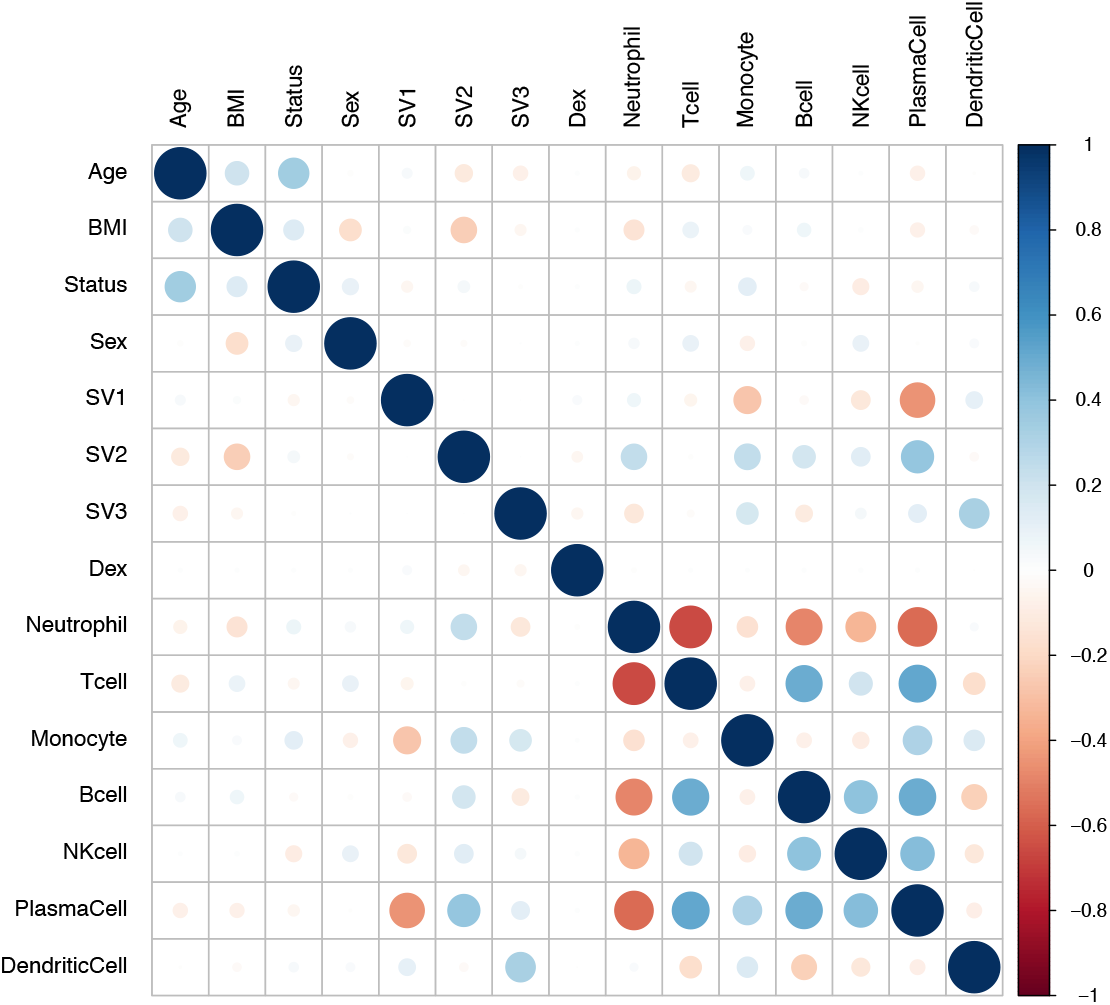
Correlation matrix of co-variants, surrogate variables (SVs) and estimated cell proportions based on CellCode.

**Figure S2:**
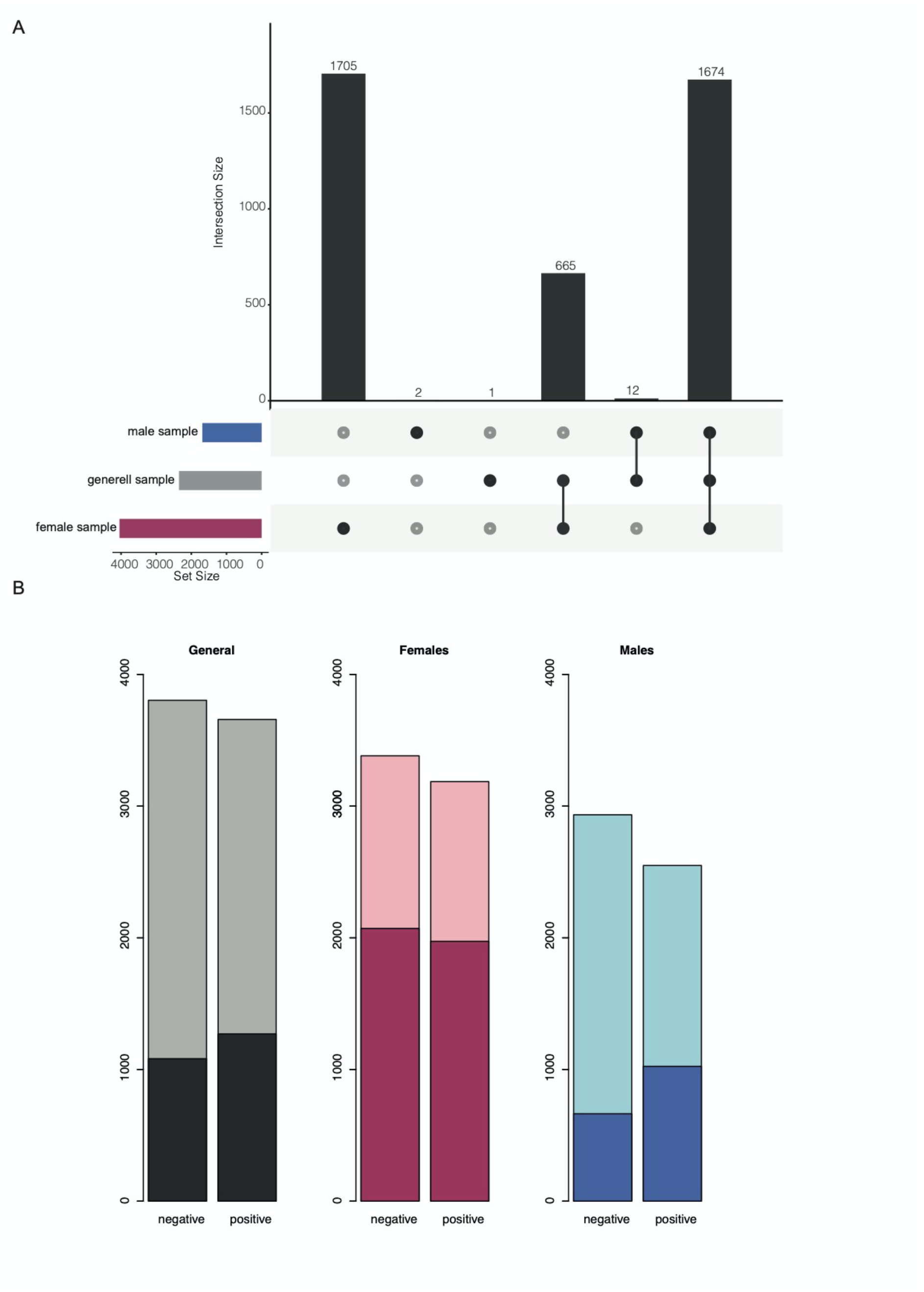
**A)** Upset plot of significant transcripts identified in combined sample, male sample, and female sample that meet absolute log_2_ FC threshold of > 0.2. **B)** Counts of significant negative and positive fold changes and significant changes surpassing an absolute log_2_ FC of 0.2 identified in the combined sample (significant gray, threshold overlaid in black), the female sample (significant pink, threshold overlaid in maroon) and the male sample (significant light blue, threshold overlaid in dark blue).

**Figure S3:**
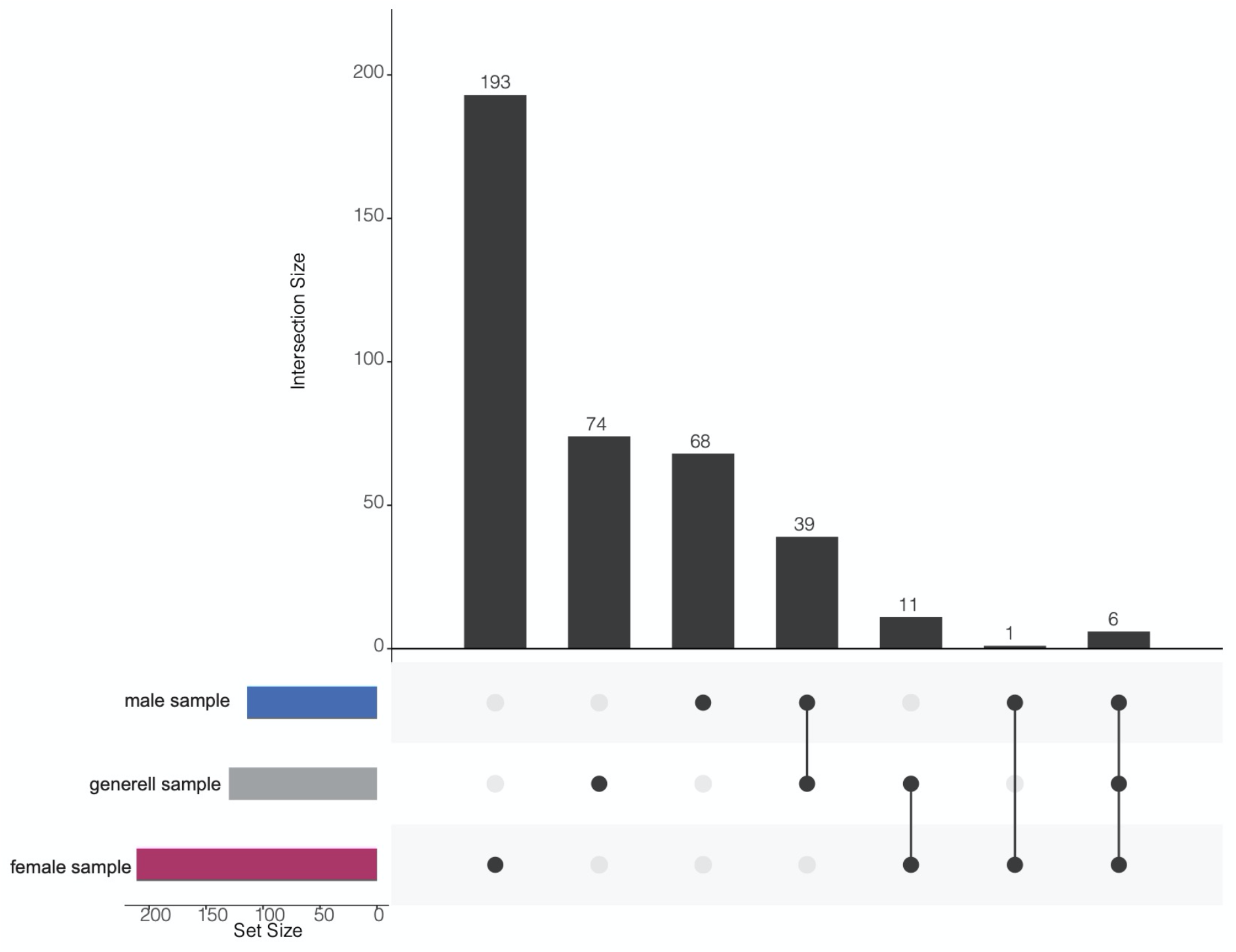
Upset plot of significant etranscripts identified in combined sample, male sample, and female sample that meet absolute log_2_ FC threshold of > 0.2.

**Figure S4:**
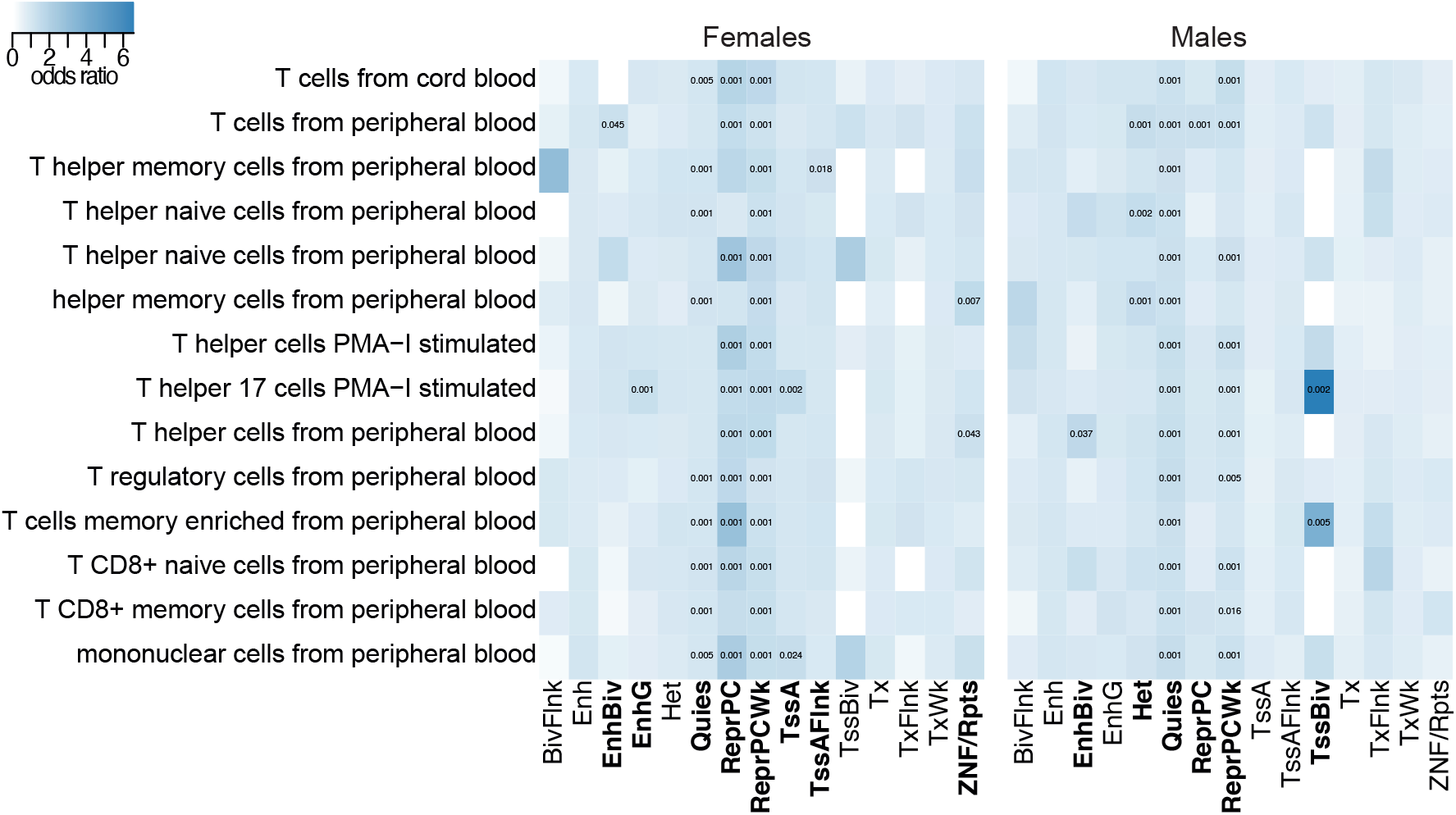
Heatmap of enrichment results for GR-response tag eSNPs and predicted ChromHMM states for sex-stratified tag eSNPs. Colors displayed indicate fold enrichment and significant permutation *p-value* are written and were derived on the basis of 1,000 random sets of baseline eSNPs matched for allele frequency and size. We observed that male and female sex-stratified GR eSNPs were significantly enriched within repressed polycomb, bivalent enhancer and quiescent states (enrichment *p-value*s < 0.05) among the tissue group of blood and T-cells (*n* = 14 cell lines). For 70% of the blood tissue group cell lines, the state for ZNF genes and repeats (*n =* 2 cell lines), genic enhancers (*n =* 1 cell line) and active transcription start site (TssA and TssAFlnk, *n =* 3 cell lines) were significantly enriched only for female GR eSNPs, see Figure 3C. Male GR eSNPs were enriched for heterochromatin (*n* = 3 cell lines) and bivalent/poised TSS (*n* = 2 cell lines).

**Figure S5:**
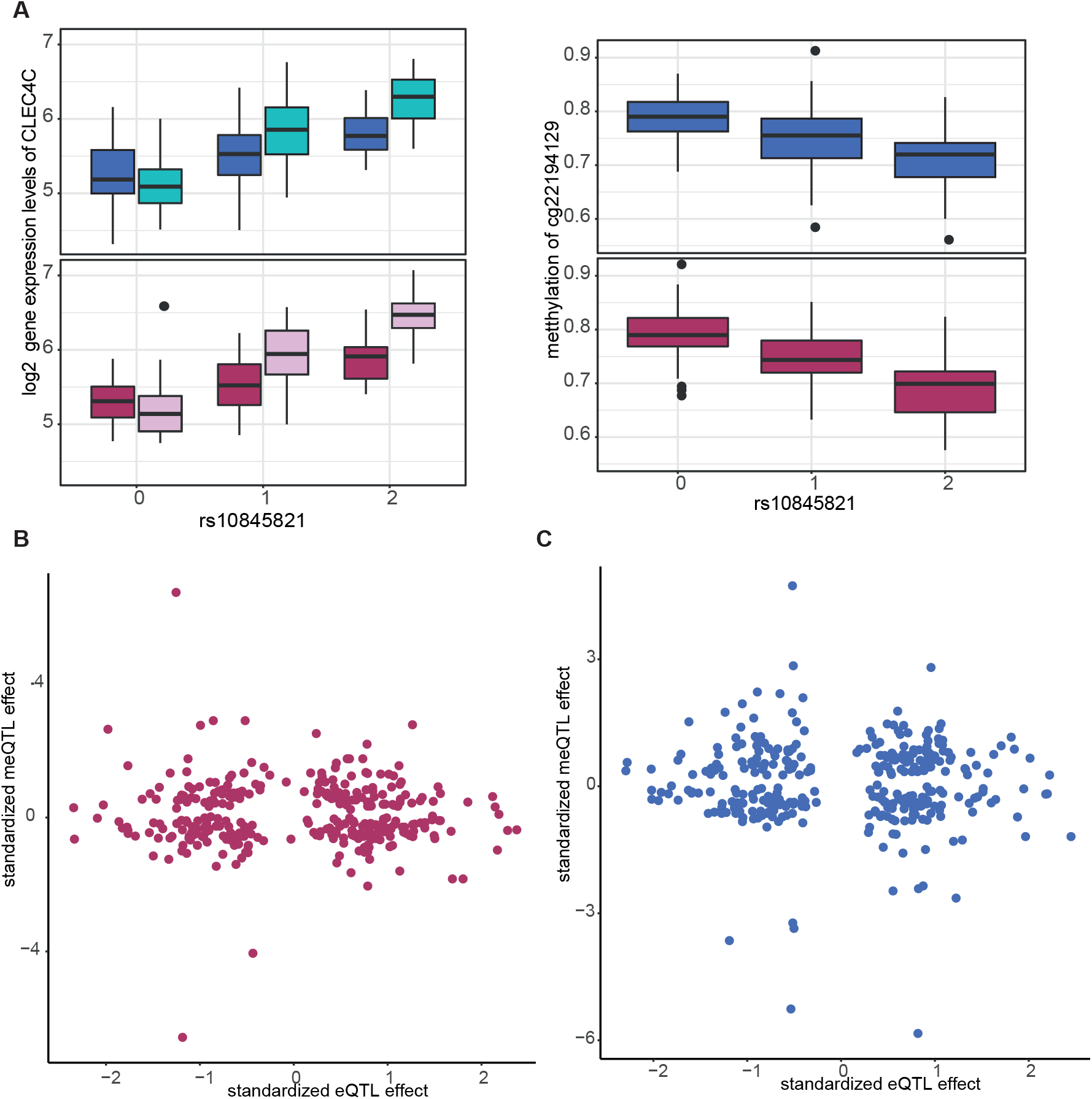
Relationships between eQTL effects and meQTL effects for **A)** females and **B)** males.

**Figure S6:**
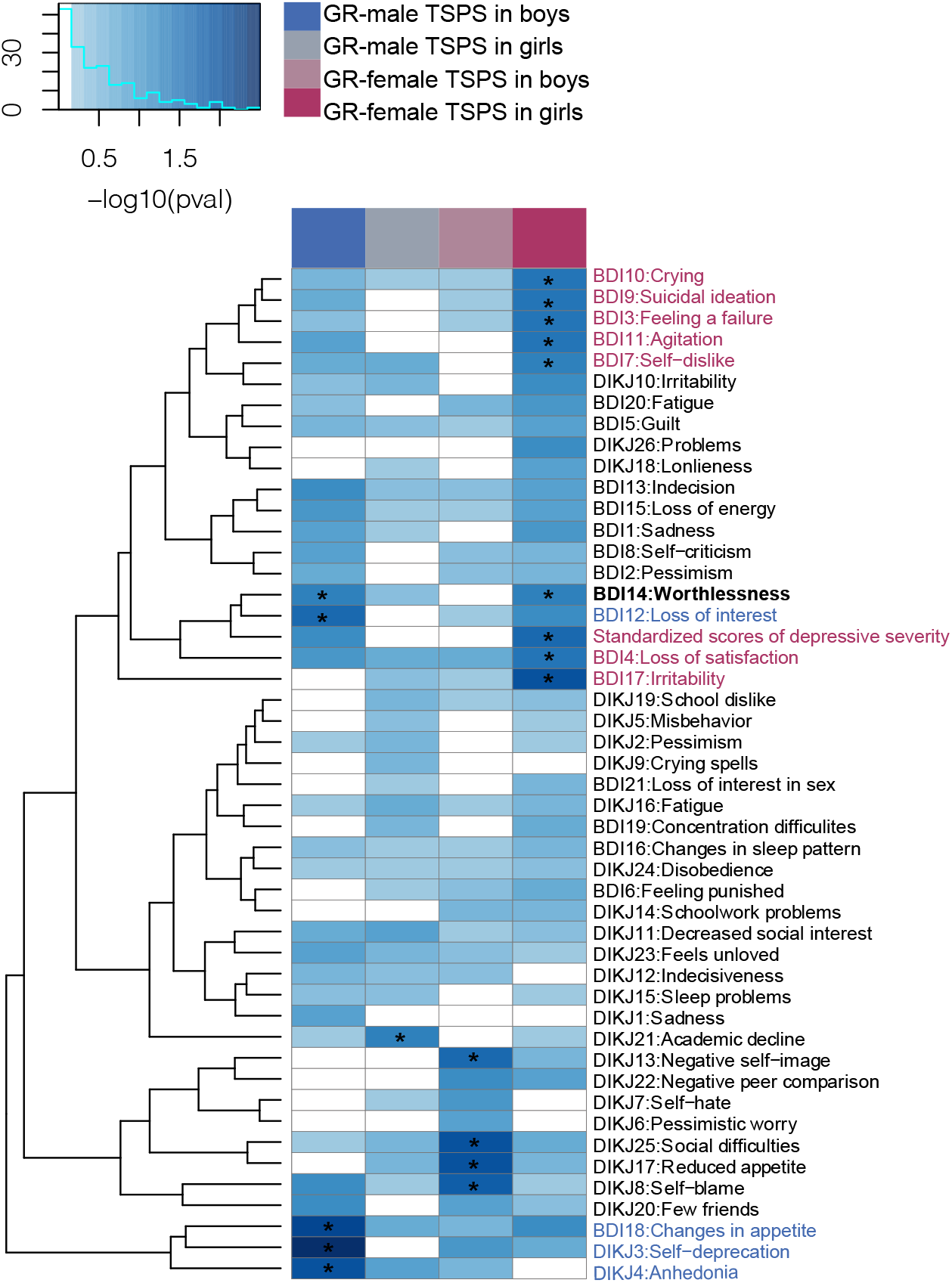
TSPS scores in correlation with individual depressive symptoms, as measured by the BDI and DIKJ.

## Supplementary Tables

**Table S1**: Analysis of sex-dependent GR-DE effects. Note. Probe_Id = Illumina probe identifier; Estimate = regression beta value; Std. Error = standard error; Pr(>|t|) = nominal *p-value*; padj = FDR-adjusted *p-value*; Chr = chromosome; opp = whether or not the effect direction is opposite for males and females.

**Table S2**: Differentially regulated transcripts across model (GR-DE transcripts, full sample, males, and females). Note. Probe_Id = Illumina probe identifier; Estimate = regression beta value; Pval = nominal *p-value* full model; Padj = FDR-adjusted *p-value*s which represent the significance of a regression model; Chr = chromosome; log2FC = log_2_ fold change; FC = fold change.

**Table S3**: List of combined analysis GR-response cis-eQTL results. Note. Probe_Id = Illumina Identifier; SNP = rsID; CHR = chromosome; BP = base pair; A1= allele 1; A2 = allele 2; Location =genomic context location; nearByGene = gene in closest proximity; P_start = starting position of the probe; P_end = ending position of the probe.

**Table S4**: List of GR-response cis-eQTL results of females. Same column labels as S3.

**Table S5**: List of GR-response cis-eQTL results of males. Same column labels as S3.

**Table S6**: Results for interaction effect of SNP and sex on etranscript gene expression. Note. Probe_Id = Illumina Identifier; Estimate = regression beta value; Std. Error = standard error; Pr(>|t|) = nominal *p-value*; AdjP = FDR-adjusted *p-value*s which represent the significance of a regression model; SNP = rsIDl; Sex = male or female eSNP set.

**Table S7**: Results of pathway analysis of the significant etranscript sets of GR-response *cis-n* eQTL identified in males and females. Note. GOBPID = the ID of biological process in GO database; Pvalue = nominal p value; ExpCount = expected number of genes in the enriched partition which map to this GO term; Count = number of genes in the enriched partition which map to this GO term; Size = number of genes within this GO Term; Term = Gene Ontology term description.

**Table S8**: List of baseline cis-eQTL results. Same column labels as S3.

**Table S9**: List of baseline cis-eQTL results of females. Same column labels as S3.

**Table S10**: List of baselines cis-eQTL results of males. Same column labels as S3. modules.

## METHODS

### Study samples

#### MPIP cohort

Participants consisted of 289 Caucasian individuals of the Max Planck Institute of Psychiatry (MPIP), 93 women and 196 men. Sex was defined by the sex chromosomes (X and Y), which is distinct from the biopsychosocial concept of gender (Davis & Stranger, 2019). Of the participants, 129 (81 men, 48 women) were being treated for MDD treated at the MPIP’s hospital in Munich and the remaining were 160 (115 men, 45 women) were healthy controls with no history of a depressive disorder, see Table 1. Recruitment strategies and further characterization of the MPIP cohort have been described previously (Arloth, Bogdan, et al., 2015). Baseline whole blood samples were obtained at 6pm after 2 hours of fasting and abstention from coffee and physical activity. Subjects then received 1.5 mg oral dexamethasone and a second blood draw was performed at 9pm three hours after dexamethasone ingestion. Plasma dexamethasone concentrations were assessed in serum samples drawn at 9pm using Liquid chromatography-tandem mass spectrometry on API4000 (AB Sciex).

### LMU cohort

The clinical LMU cohort consists if 584 Caucasian children and adolescents (ages 7–18 years old) recruited from two child and adolescent clinics in Munich: 350 cases with a current diagnosis or history of major depression and 234 healthy control subjects with no history of a psychiatric disorder. The presence or absence of depression was determined by a well-established diagnostic interview (Adornetto et al., 2008). Further characterization of the cohort and psychometric measures are described in (Halldorsdottir et al., 2019) and Table 1. To assess the severity of depressive symptoms, the Children’s Depression Inventory (CDI) was administered to youths ≤12 years old, and the Beck Depression Inventory–II (BDI-II) was administered to participants >12 years old. Scores from the CDI and the BDI-II were standardized using z scores to perform the analyses on the whole sample. We explored potential sex differences in trauma exposure and did not find evidence of significant sex differences in history of sexual abuse or overall stress exposure levels.

### recMDD cohort

The recMDD cohort consisted of 1,774 Caucasian individuals recruited at the MPIP in Munich, Germany and two satellite hospitals in the Munich metropolitan area (BKH Augsburg and Klinikum Ingolstadt): 756 controls and 879 cases diagnosed with recurrent major depression. Please see (Muglia et al., 2010) for more details on sample recruitment and characterization and Table 1. A subset of *n =* 567 individuals was used in this manuscript.

All studies were approved by the local ethics committees and were conducted in accordance with the current version of the Declaration of Helsinki.

### Gene expression data

Whole blood RNA (Baseline and GR-response) from the MPIP cohort samples was collected using PAXgene Blood RNA Tubes (PreAnalytiX) and processed as described previously (Arloth, Bader, et al., 2015). The RNA was then hybridized to Illumina HT-12 v3 and v4 expression Bead Chips (Illumina, San Diego, CA). Raw probe intensities were exported using Illumina’s GenomeStudio and further statistical processing was carried out using R version 3.2.1. All 29,075 probes present on both BeadChips (v3 vs. v4), excluding X and Y chromosomes as well as cross-hybridizing probes identified by using the Re-Annotator pipeline (Arloth, Bader, et al., 2015) were first filtered with a detection *p-value* of 0.05 in at least 50% of the samples, leaving 11,994 autosomal expression array probes. Subsequently, each probe was transformed and normalized through variance stabilization and normalization (VSN) (Johnson et al., 2007). Technical batch effects were identified by inspecting the association of the first principal components of the expression levels for all known batch effects and then adjusted using ComBat (Johnson et al., 2007) with slide, amplification round, array version, and amplification plate column as fixed effects. The position of the gene expression probe and gene symbols were annotated using the Re-Annotator pipeline (Arloth, Bader, et al., 2015) based on GRCh37 (hg19) RefSeq (Pruitt et al., 2012). Surrogate Variable Analysis (SVA) (Leek et al., 2012) was used to account for confounding as a result of batch effects, cell proportion or unknown factors using the SVA package in Bioconductor version 3.3. We compared the significant SVs to the estimated fractions of different blood cell types derived from the residuals of the transcriptome-wide gene expression values using CellCODE (Chikina et al., 2015), see **Supplementary Figure 1** for the SV correlations with blood cell count and known confounding factors. The log FC of gene expression was calculated as the difference in gene expression between post dexamethasone and baseline standardized to baseline.

### Genotype data and Imputation

Genotype data was generated for each cohort individually. Human DNA of the MPIP cohort samples was isolated from EDTA blood samples using the Gentra Puregene Blood Kit (Qiagen) with standardized protocols. Genome-wide SNP genotyping was performed using Illumina Human610-Quad (*n =* 173) and OmniExpress (*n =* 120) genotyping BeadChips according to the manufacturer’s standard protocols. recMDD cohort samples have been genotype on the Illumina-550 BeadChip and details on the genotyping methods have been previously published (Muglia et al., 2010). Quality control was conducted in PLINK 1.90b3s (Chang et al., 2015) or higher for each cohort and genotyping BeadChip separately. QC steps on samples included removal of individuals with a missing rate >2%, cryptic relatives (*PI-HAT* >0.0125), an autosomal heterozygosity deviation (|*F*_*he*t_| >4 SD), and genetic outliers (distance in the ancestry components from the mean >4 SD). QC steps on variants included removal of variants with a call rate <98%, a MAF <1%, and HWE test *p-value*s ≤10^−6^.

Furthermore, variants on non-autosomal chromosomes were excluded. Imputation was performed separately for each cohort and genotyping BeadChip with IMPUTE2, following phasing in SHAPEIT, using the 1,000 genomes phase I reference panel (released in June 2014, all samples). QC of imputed probabilities was conducted in QCTOOL 1.4. Imputed SNPs were excluded if MAF <1%, HWE test *p-value*s ≤10^−6^, or an INFO metric <0.8. SNP coordinates are given according to hg19. SNPs were further processed in PLINK and variants were excluded if their MAF < 5%.

Genotyping of the LMU cohort was performed with the Infinium Global Screening Array BeadChip. Genotyping of the recMDD was performed with Illumina Human610-Quad BeadChips. Further detail on the genotyping and imputation methods used can be found in the individual papers LMU: (Halldorsdottir et al., 2019) and recMDD: (Muglia et al., 2010).

### Differential gene expression analysis (DEA)

To observe both dexamethasone-dependent changes in gene expression, and sex-stratified effects of dexamethasone, we ran the following models. First, we calculated the effect of sex on the difference in gene expression between baseline and post dexamethasone controlling for age, BMI, depression status, and cell type.

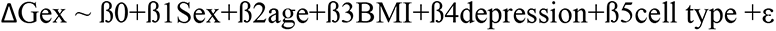

Second, a main effects linear model isolates the probes that are regulated by dexamethasone administration, controlling for sex, age, BMI, depression status, and cell type. Finally, the same main effect linear model was ran separately in males and females (not controlling for sex).

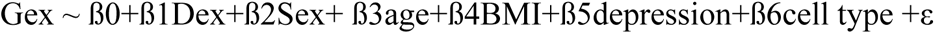

### Expression quantitative trait loci analysis

The eQTL analysis was restricted to those SNPs within 1Mb upstream or downstream For each gene expression array probe a linear model of the log fold change on gene expression was constructed between baseline and GR-response standardized to baseline and gender (only for the combined analysis). The residuals from the linear regression were used as phenotype values in the following analyses. PLINK v2 (Chang et al., 2015) was used to test for *cis-*association between all imputed SNPs and transcriptional response as previously described (Arloth et al., 2015).

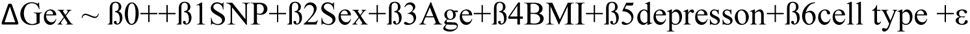

We ran the same model, but separated for males and females for the sex-biased eQTL analysis.

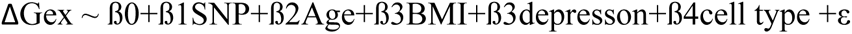

Finally, for each set of sex-stratified etranscript gene expression array probe (identified by the models ran separately for males and females), the delta value between dexamethasone and baseline was predicted by the interaction of sex and eSNP, controlling for age, BMI, disease-state and SVAs.

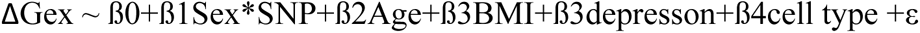

As eQTL data were composed of two kinds of data: genotyping and expression data, we used two stages of multiple testing correction: (i) SNP level correction: for each *cis*-region (array probe) we performed a permutation test. The sample identifiers in the gene expression data were shuffled in order to preserve the structure in the genotype data (LD). A total of 500,000 permutations were carried out per probe and the empirical *P* values were adjusted using the Westfall-Young correction for the number of SNPs per probe, i.e., maxT procedure of Westfall-Young (Terada et al., 2013). (ii) Probe level correction: *cis*-regions with an extensive LD structure will increase the number of false positive eQTLs (Westra et al., 2013). Therefore, we applied the Benjamini-Hochberg method to correct the maxT adjusted *P* value significance by using only the most significant and independent SNPs per probe (tag SNPs). The number of tag eSNPs per *cis*-region was identified by LD pruning and “clumping” the SNPs using the “clump” command in PLINK (using distance < 1Mb and *r*^*2*^ ≤ 0.2 as setting). Each tag SNP forms a SNP bin, by aggregating all other SNPs into bins by tag SNP at *r*^*2*^ ≤ 0.2 and distance < 1Mb, such that all SNPs within a given bin were correlated to their corresponding tag SNP, but not to any other tag SNP. We limited the false-positive SNP-probe pairs to less than 5% and therefore we considered the FDR analogue of the *P* value (*Q* value) < 5% as statistically significant.

### Power analysis

Given our different sample sizes of males and females, we determined our power for sex-stratified eQTL analyses. Given an effect size of the top eQTL for each analysis, we had 98% power in males, 57% power for females, and 79% for the combined sample with 0.07, 0.04, 0.02 as regression coefficients. For adequate power in the female only sample, we estimated that a sample of 382 would be required for equal power to the male analysis (98%) to detect *cis*-eQTLs. Power estimates were calculated using the G*power 3.19.4 application (Faul et al., 2007).

### Pathway analysis

The Bioconductor package TheGOstats was used to explore the gene ontologies of groups of transcripts over represented relative to all transcripts explored (*n* = 11,272 probes after quality control, or the gene ‘universe’). In terms of ontologies, we tested for biological processes and used the human genome wide annotation (org.Hs.eg.db). Due to high dependencies among GO terms, nominal *p-values* are reported. For descriptive purposes, the top gene ontologies were selected in the analysis of etranscripts overlapping with transcripts identified in BA25 in association with MDD.

### Genomic region annotation

eSNPs were overlapped with genomic annotation from UCSC for the hg19 genome build using *TxDb*.*Hsapiens*.*UCSC*.*hg19*.*knownGene* and *ChIPseeker* Bioconductor *R* packages.

### Epigenetic enrichment analysis

To identify whether GR-response eSNPs were enriched for GR binding sites or co-localize with specific chromatin states, we used the Encode NR3C1 ChIP-seq data from GM12878 LCLs treated with dexamethasone (accession: GSE45638) and the 15-state ChromHMM (Ernst & Kellis, 2017) annotation of the Roadmap Epigenomics project among all cell lines of the blood and T-cell tissue group (*n =* 14 cell lines). We calculated the position-based overlap of the GR-response tag eSNPs and chromatin states for gender separately and compared the overlap observed with 1,000 equal sized sets of baseline tag eSNPs adjusting for MAF. We used DeepSEA, a deep neural network pretrained with DNase-seq and ChIP-seq data from the ENCODE project, to predict the likelihood that GR-sex eSNPs exert regulatory effects on chromatin features comparing the reference to alternative SNP.

The coordinates of AR and ER binding sites were downloaded from Remap. There was no enrichment of sex-biased eSNPs for sex hormone receptors beyond baseline sex-biased eSNPs. To test for enrichment of TFs, we used the R package ReMapEnrich () using the 2018 Remap catalog on hg19.

We annotated the eSNPs using DeepSEA (Zhou & Troyanskaya, 2015). DeepSEA, a deep neural network pretrained with DNase-seq and ChIP-seq data from the ENCODE project, predicts the presence of histone marks, DNase hypersensitive regions (DHS) or TF binding for a given 1 kb sequence. The likelihood that a specific genetic variant influences regulatory chromatin features is estimated by comparing predicted probabilities of two sequences where the bases at the central position are the reference and alternative alleles of a given variant.

### DNA methylation data and meQTL analysis

For a subset of the reCMDD cohort (*n =* 567 individuals), genomic DNA was extracted from whole blood using the Gentra Puregene Blood Kit (QIAGEN). DNA quality and quantity of both was assessed with the NanoDrop 2000 Spectrophotometer (Thermo Scientific) and Quant-iT Picogreen (Invitrogen). Genomic DNA was bisulfite converted using the Zymo EZ-96 DNA Methylation Kit (Zymo Research) and DNA methylation levels were assessed for >480,000 CpG sites using the Illumina HumanMethylation450K BeadChips. Hybridization and processing were performed according to the manufacturer’s instructions. QC of methylation data, including intensity readouts, filtering (detection *p-value* >0.01 in at least 75% of the samples), cellular composition estimated using *FlowSorted*.*Blood*.*450k* data and “estimateCellCounts” function, as well as beta calculation (“getBeta” function) were done using the *minfi* Bioconductor *R* package. CpG sites on sex chromosomes, CpG site probes found to have SNPs at the CpG site itself or in the single-base extension site with a MAF ≥1% in the 1,000 genomes project EUR population and non-specific binding CpG site probes according to (Chen et al., 2013) were removed. We performed a re-alignment of the CpG site probe sequences using *Bismark*. This yielded 425,883 CpG sites for further analysis. The data were then normalized using functional normalization (“preprocessFunnorm” function in *minfi*) (Aryee et al., 2014). Technical batch effects were identified by inspecting the association of the first principal components of the methylation levels with plate and plate position. The data were then adjusted using “*ComBat”* function of the Bioconductor *R* package *sva*. CpG coordinates are given according to hg19.

For the meQTL analysis, linear regression models were fit for males and females separately and for each CpG site to test the relationship between the whole blood DNA methylation (beta values) and proximal SNP genotype (in dosage format) within 1Mb up- or downstream of the SNP using the *R* package *MatrixEQTL* (Shabalin, 2012), in order to detect *cis*-meQTLs. Blood cell counts and age were included as covariates. Significance after multiple testing was adjusted using a false discovery rate (FDR) of 5%.

### Enrichment in Labonté et al., 2017

To test for enrichment of male and female GR-DE transcripts within male and female MDD transcriptional patterns in six brain regions, we used the ‘GeneOverlap’ R package to determine the significance of overlap from two lists based on the Jaccard index, given the size of common genes tested in the two data sets (n = 8,683 genes). Enrichment for male and female GR eQTL associated etranscripts was tested in comparison to the overlap observed for baseline GR eQTL associated etranscripts based on odds ratios and p values from the Fisher’s exact test.

### GWAS enrichment analysis

The nominal GWAS results *p-value* < 0.05 of the Psychiatric Genomics Consortium (PGC) for different psychiatric disorders: schizophrenia (SCZ2) (Ripke et al., 2014), bipolar disorder (BIP) (Stahl et al., 2019), MDD (MDD1-3) (Howard et al., 2019; Ripke et al., 2013; Wray et al., 2018b), autism spectrum disorder (AUT) (Consortium, 2017), attention-deficit/hyperactivity disorder (ADHD) (Demontis et al., 2019), PTSD (Nievergelt et al., 2019), Tourette syndrome (TS) (Yu et al., 2019) and cross disorder (CDG1&2) (P. H. Lee et al., 2019; Smoller et al., 2013) and non-psychiatric phenotypes: the Social Science Genetic Association Consortium (SSGAC) for educational attainment (EA) (Lee et al., 2018), cannabis use (Pasman et al., 2018), Type 2 diabetes (T2D) (Xue et al., 2018) and the Complex Trait Genetics Lab of the VU University of Amsterdam for intelligence (Savage et al., 2018) were used for comparison with our GR-response results. Thereby the overlap between the tag SNPs comprised in our eQTL bins and the SNPs identified by these studies were calculated. The enrichment eQTL-SNPs and GWAS risk-SNPs was tested in comparison with 1,000 MAF-matched baseline tag eSNP sets.

### Transcriptional sensitivity profile score (TSPS)

TSPSs were based on the sets of significant GR-response tag eSNPs for males and females in the independent clinical LMU cohort. Of the 601 female GR-response eSNPs, 562 were available in the test cohort (with 57 proxy SNPs, r^2^>0.6), and of the 668 male, 650 (with 47 proxy SNPs, r^2^>0.6) used for calculation of the TSPS. Risk alleles were determined by the coefficient from the GR-response eQTL analysis, in such that the alleles associated with higher absolute coefficients were coded as a risk allele. Absolute coefficient from the eQTL calculation were further included as weights. The scores were corrected for the number of SNPs. For eSNPs regulating multiple transcripts, we included each eQTL association and their beta coefficient. A higher TSPS thus denotes a larger number of alleles associated with larger GR-induced transcriptional response.

## Declarations

### Availability of data and material

Data from MPIP gene expression experiment are deposited at the GEO repository under GEO: GSE64930 and recMDD methylation under: GSE125105. Data analysis code is available at https://github.com/jArloth/sex-specific-GR-response-Analyses.

### Authors’ approval and contributions

All authors have seen and approved this manuscript for submission.

Conception and design: Janine Arloth acquisition of MPIP data: Elisabeth B. Binder acquisition of recMDD data: Susanne Lucae, Bertram Müller-Myhsok acquisition of LMU data: Charlotte Piechaczek, Lisa Feldmann, Franz Joseph Freisleder, Ellen Greimel, Gerd Schulte-Körne Formal analysis: Sarah Moore, Janine Arloth analysis and interpretation of data: Sarah Moore, Thorhildur Halldorsdottir, Jade Martins, Nikola S.Müller, Elisabeth B. Binder, Janine Arloth drafting or revising the manuscript: Sarah Moore, Thorhildur Halldorsdottir, Elisabeth B. Binder, Janine Arloth

## Acknowledgements

We thank all individuals who agreed to participate in and provided blood samples for this study. We would like to thank Rick Jansen, Darina Czamara, Richa Batra, Linda Krause, Christoph Orgis, Karolina Worf, and Yue Hu for useful discussions on the approach.

## Ethics approval and consent to participate

All studies were approved by the local ethics committee and all individuals gave written informed consent. All experimental methods comply with the Helsinki Declaration.

## Supplementary Material

### Analysis of sex-biased GR-DE effects

There may be transcripts which demonstrate a sex-biased effect of dexamethasone, which we tested by modeling sex as a predictor of the difference in gene expression between baseline and post dexamethasone administration standardized by baseline expression (see Methods), previously defined as GR-response values (Arloth et al., 2015). In this model, 26 transcripts demonstrated a significant effect of sex on dexamethasone change after multiple test correction (FDR< 0.05 **Supplementary Table 1**, and the majority (*n* = 15 transcripts; 57.69%) were already identified by the main effect model above or by the models stratified by sex. Across the 26 significant hits, the fold changes surpassed an absolute threshold of 0.2 for 7 transcripts, in which males and females demonstrated consistent directions of effects (i.e., one sex demonstrate a stronger effect than the other). The direction of the effect between baseline and dexamethasone was opposite for females compared to males for only two transcripts, but these fold changes did not surpass the fold change cut off for males or for females (see **Supplementary Table 1**).

Given our different sample sizes of males (*n* = 196) and females (*n* = 93), we tested the robustness of our results by using down-sampled sets of males matching female same size. We randomly drew 93 males from the full male sample to assess the number of significant transcripts found across 100 permutations. We found that the range of significant GR-DE transcripts found in males was between 3,125 and 3,251, with a mean of 3,178.2 ±25, relative to 5,438 DE-GR transcripts identified with the full male sample and 6,568 DE-GR transcripts identified in females. All permutations yielded less transcripts than identified in the female sample of equal sample size.

### Effect size filtered transcripts and etranscripts

**Supplementary Figure 2A** shows how these overlapping results across the general sample, males, and females with results are filtered by absolute log2 fold changes (> 0.2). A large proportion of the transcripts that were found by the main model to be significantly regulated by dexamethasone only surpassed the log2 fold change cut off in females (42.2%, *n* = 1705; shown in the shift to a larger number of transcripts unique to females in **Supplementary Figure 2A** relative to **Figure 1C**). In contrast, only 2 (0.12%) significant transcripts found in the main model surpassed the cut off uniquely in males. A minority of significant transcripts, 1,674 met the cut off across models. Taken together, these comparisons show that many of the transcriptional changes induced by dexamethasone identified in the main model were driven by larger effects in females. **Supplementary Figure 2B** similarly shows this trend, with a larger proportion of significant transcripts reaching the log2 fold change cut off from the female model relative to the general or male models.

We next assessed the size of the effects of those etranscripts that overlap with the transcripts from the differential expression analysis. Out of the 324 overlapping male etranscripts, 114 (35%) surpassed the absolute fold change threshold of 0.2 (range: -0.35 to 0.75), and 210 out of 333 (63%) of the overlapping female etranscripts surpassed the log2 fold change cutoff (range: -0.52 to 1.5). Thus, consistent with the GR-DE analysis, overlapping etranscripts in females exhibited larger fold changes relative to males (Wilcoxon p-value = 0.05).

Relative to the differential expression analysis, etranscripts meeting the fold change cut off were again largely found in males or females independently rather than the general, across sex analysis. Specifically, only six effect size filtered etranscripts were found in the general model and in males and females independently (**Supplementary Figure 3**).

### Baseline eQTLs

When analyzing only baseline expression we found 4,202 general baseline eQTL bins (5,722 tag eSNPs of 167,885 correlated eSNPs and 2,909 gene expression probes and 2,550 genes, **Supplementary Table 8**). These results were compared to a larger publicly available database-the Biobank-Based Integrative Omics Study (BIOS) (>2,000 whole blood samples) and whole blood eQTLs from the Genotype-Tissue Expression project v8 (GTEx; 670 donors) (Consortium, 2020; Zhernakova et al., 2017). We found that 87% of our baseline eQTL genes overlapped with significant BIOS and 92% with significant GTEx eQTL genes.

We assessed male and female eQTLs separately at baseline and aimed to validate these baseline sex-biased eQTLs in existing data sets. We identified two times the number of eQTLs for males then for females, e.g. 1,960 eQTL bins (1,561 gene expression probes and 1,638 tag eSNPs; **Supplementary Table 9**) in females and 4,433 eQTL bins (2,561 gene expression probes and 3,209 tag eSNPs; **Supplementary Table 10**) in males only. Of note, for baseline eQTLs, the effects were significantly smaller for males relative to females (Wilcoxon p-value = 0.02). We found that 86% of our baseline male eQTL genes overlapped with nominal significant GTEx male eQTL genes (*p-value* < 0.05) and 84% of our baseline female eQTLs with nominal significant GTEx female eQTL genes. Thus, the sex differences we observed in eQTLs in the blood transcriptome at baseline are consistent with the extant literature, supporting the accuracy of identified sex differences.

## References

Abel, K. M., Drake, R., & Goldstein, J. M. (2010). Sex differences in schizophrenia. International Review of Psychiatry, 22(5), 417–428. https://doi.org/10.3109/09540261.2010.515205

Adornetto, C., Hensdiek, M., Meyer, A., In-Albon, T., Federer, M., & Schneider, S. (2008). The factor structure of the Childhood Anxiety Sensitivity Index in German children. Journal of Behavior Therapy and Experimental Psychiatry, 39(4), 404–416. https://doi.org/10.1016/j.jbtep.2008.01.001

Aguet, F., Barbeira, A. N., Bonazzola, R., Brown, A., Castel, S. E., Jo, B., Kasela, S., Kim-Hellmuth, S., Liang, Y., Oliva, M., Flynn, E. D., Parsana, P., Fresard, L., Gamazon, E. R., Hamel, A. R., He, Y., Hormozdiari, F., Mohammadi, P., Muñoz-Aguirre, M., … Volpi, S. (2020a). The impact of sex on gene expression across human tissues. Science, 369(6509). https://doi.org/10.1126/SCIENCE.ABA3066

Aguet, F., Barbeira, A. N., Bonazzola, R., Brown, A., Castel, S. E., Jo, B., Kasela, S., Kim-Hellmuth, S., Liang, Y., Oliva, M., Flynn, E. D., Parsana, P., Fresard, L., Gamazon, E. R., Hamel, A. R., He, Y., Hormozdiari, F., Mohammadi, P., Muñoz-Aguirre, M., … Volpi, S. (2020b). The impact of sex on gene expression across human tissues. Science, 369(6509). https://doi.org/10.1126/SCIENCE.ABA3066

Arloth, J., Bader, D. M., Röh, S., & Altmann, A. (2015). Re-Annotator: Annotation pipeline for microarray probe sequences. PLoS ONE, 10(10). https://doi.org/10.1371/journal.pone.0139516

Arloth, J., Bogdan, R., Weber, P., Frishman, G., Menke, A., Wagner, K. V., Balsevich, G., Schmidt, M. V., Karbalai, N., Czamara, D., Altmann, A., Trümbach, D., Wurst, W., Mehta, D., Uhr, M., Klengel, T., Erhardt, A., Carey, C. E., Conley, E. D., … Binder, E. B. (2015). Genetic Differences in the Immediate Transcriptome Response to Stress Predict Risk-Related Brain Function and Psychiatric Disorders. Neuron, 86(5), 1189–1202. https://doi.org/10.1016/j.neuron.2015.05.034

Aryee, M. J., Jaffe, A. E., Corrada-Bravo, H., Ladd-Acosta, C., Feinberg, A. P., Hansen, K. D., & Irizarry, R. A. (2014). Minfi: A flexible and comprehensive Bioconductor package for the analysis of Infinium DNA methylation microarrays. Bioinformatics, 30(10), 1363–1369. https://doi.org/10.1093/bioinformatics/btu049

Bale, T. L., & Epperson, C. N. (2015). Sex differences and stress across the lifespan. Nature Neuroscience, 18(10), 1413–1420. https://doi.org/10.1038/nn.4112

Bekhbat, M., & Neigh, G. N. (2018). Sex differences in the neuro-immune consequences of stress: Focus on depression and anxiety. Brain, Behavior, and Immunity, 67, 1–12. https://doi.org/10.1016/J.BBI.2017.02.006

Bezchlibnyk, Y. B., Cheng, J., Bijanki, K. R., Mayberg, H. S., & Gross, R. E. (2018). Subgenual Cingulate Deep Brain Stimulation for Treatment-Resistant Depression. In Neuromodulation (pp. 1099–1118). Elsevier. https://doi.org/10.1016/b978-0-12-805353-9.00091-7

Boraska, V., Jerončić, A., Colonna, V., Southam, L., Nyholt, D. R., William Rayner, N., Perry, J. R. B., Toniolo, D., Albrecht, E., Ang, W., Bandinelli, S., Barbalic, M., Barroso, I., Beckmann, J. S., Biffar, R., Boomsma, D., Campbell, H., Corre, T., Erdmann, J., … Zeggini, E. (2012). Genome-wide meta-analysis of common variant differences between men and women. Human Molecular Genetics, 21(21), 4805–4815. https://doi.org/10.1093/hmg/dds304

Boyd, A., Van de Velde, S., Vilagut, G., de Graaf, R., O׳Neill, S., Florescu, S., Alonso, J., Kovess-Masfety, V., & EU-WMH Investigators. (2015). Gender differences in mental disorders and suicidality in Europe: Results from a large cross-sectional population-based study. Journal of Affective Disorders, 173, 245–254. https://doi.org/10.1016/j.jad.2014.11.002

Brivio, E., Lopez, J. P., & Chen, A. (2020). Sex differences: Transcriptional signatures of stress exposure in male and female brains. In Genes, Brain and Behavior (Vol. 19, Issue 3). Blackwell Publishing Ltd. https://doi.org/10.1111/gbb.12643

Chadwick, L. H. (2012). The NIH Roadmap Epigenomics Program data resource. In Epigenomics (Vol. 4, Issue 3, pp. 317–324). https://doi.org/10.2217/epi.12.18

Chang, C. C., Chow, C. C., Tellier, L. C. A. M., Vattikuti, S., Purcell, S. M., & Lee, J. J. (2015). Second-generation PLINK: Rising to the challenge of larger and richer datasets. GigaScience, 4(1). https://doi.org/10.1186/s13742-015-0047-8

Chen, Y., Lemire, M., Choufani, S., Butcher, D. T., Grafodatskaya, D., Zanke, B. W., Gallinger, S., Hudson, T. J., & Weksberg, R. (2013). Discovery of cross-reactive probes and polymorphic CpGs in the Illumina Infinium HumanMethylation450 microarray. Epigenetics, 8(2), 203–209. https://doi.org/10.4161/epi.23470

Chikina, M., Zaslavsky, E., & Sealfon, S. C. (2015). CellCODE: A robust latent variable approach to differential expression analysis for heterogeneous cell populations. Bioinformatics, 31(10), 1584–1591. https://doi.org/10.1093/bioinformatics/btv015

Childs, E., Dlugos, A., & De Wit, H. (2010). Cardiovascular, hormonal, and emotional responses to the TSST in relation to sex and menstrual cycle phase. Psychophysiology, 47(3), 550–559. https://doi.org/10.1111/j.1469-8986.2009.00961.x

Consortium, A. S. D. W. G. of T. P. G. (2017). Meta-analysis of GWAS of over 16,000 individuals with autism spectrum disorder highlights a novel locus at 10q24.32 and a significant overlap with schizophrenia. Molecular Autism, 8(1), 21. https://doi.org/10.1186/s13229-017-0137-9

Cross-Disorder Group of the Psychiatric Genomics Consortium, Lee, P. H., Anttila, V., Won, H., Feng, Y.-C. A., Rosenthal, J., Zhu, Z., Tucker-Drob, E. M., Nivard, M. G., Grotzinger, A. D., Posthuma, D., Wang, M. M.-J., Yu, D., Stahl, E., Walters, R. K., Anney, R. J. L., Duncan, L. E., Belangero, S., Luykx, J., … Smoller, J. W. (2019). Genome wide meta-analysis identifies genomic relationships, novel loci, and pleiotropic mechanisms across eight psychiatric disorders. BioRxiv, 528117. https://doi.org/10.1101/528117

Davis, L. K., & Stranger, B. E. (2019). The new science of sex differences in neuropsychiatric traits. American Journal of Medical Genetics Part B: Neuropsychiatric Genetics, 180(6), 333–334. https://doi.org/10.1002/ajmg.b.32747

de Kloet, E. R., Joëls, M., & Holsboer, F. (2005). Stress and the brain: from adaptation to disease. Nature Reviews Neuroscience, 6(6), 463–475. https://doi.org/10.1038/nrn1683

Demontis, D., Walters, R. K., Martin, J., Mattheisen, M., Als, T. D., Agerbo, E., Baldursson, G., Belliveau, R., Bybjerg-Grauholm, J., Bækvad-Hansen, M., Cerrato, F., Chambert, K., Churchhouse, C., Dumont, A., Eriksson, N., Gandal, M., Goldstein, J. I., Grasby, K. L., Grove, J., … Neale, B. M. (2019). Discovery of the first genome-wide significant risk loci for attention deficit/hyperactivity disorder. Nature Genetics, 51(1), 63–75. https://doi.org/10.1038/s41588-018-0269-7

Diflorio, A., & Jones, I. (2010). Is sex important? Gender differences in bipolar disorder. International Review of Psychiatry, 22(5), 437–452. https://doi.org/10.3109/09540261.2010.514601

Dimas, A. S., Nica, A. C., Montgomery, S. B., Stranger, B. E., Raj, T., Buil, A., Giger, T., Lappalainen, T., Gutierrez-Arcelus, M., McCarthy, M. I., & Dermitzakis, E. T. (2012). Sex-biased genetic effects on gene regulation in humans. Genome Research, 22(12), 2368–2375. https://doi.org/10.1101/gr.134981.111

Elbau, I. G., Brücklmeier, B., Uhr, M., Arloth, J., Czamara, D., Spoormaker, V. I., Czisch, M., Stephan, K. E., Binder, E. B., & Sämann, P. G. (2018). The brain’s hemodynamic response function rapidly changes under acute psychosocial stress in association with genetic and endocrine stress response markers. Proceedings of the National Academy of Sciences of the United States of America, 115(43), E10206–E10215. https://doi.org/10.1073/pnas.1804340115

Ellegren, H., & Parsch, J. (2007). The evolution of sex-biased genes and sex-biased gene expression. Nature Reviews Genetics, 8(9), 689–698. https://doi.org/10.1038/nrg2167

Ernst, J., & Kellis, M. (2017). Chromatin-state discovery and genome annotation with ChromHMM. Nature Protocols, 12(12), 2478–2492. https://doi.org/10.1038/nprot.2017.124

Fadason, T., Schierding, W., Lumley, T., & O’Sullivan, J. M. (2018). Chromatin interactions and expression quantitative trait loci reveal genetic drivers of multimorbidities. Nature Communications, 9(1). https://doi.org/10.1038/s41467-018-07692-y

Faul, F., Erdfelder, E., Lang, A. G., & Buchner, A. (2007). G*Power 3: A flexible statistical power analysis program for the social, behavioral, and biomedical sciences. Behavior Research Methods, 39(2), 175–191. https://doi.org/10.3758/BF03193146

Gershoni, M., & Pietrokovski, S. (2017). The landscape of sex-differential transcriptome and its consequent selection in human adults. BMC Biology, 15(1). https://doi.org/10.1186/s12915-017-0352-z

Gilks, W. P., Abbott, J. K., & Morrow, E. H. (2014). Sex differences in disease genetics: evidence, evolution, and detection. Trends in Genetics, 30(10), 453–463. https://doi.org/10.1016/j.tig.2014.08.006

Gold, P. W. (2015). The organization of the stress system and its dysregulation in depressive illness. Molecular Psychiatry, 20(1), 32–47. https://doi.org/10.1038/mp.2014.163

Halldorsdottir, T., Piechaczek, C., Soares de Matos, A.P., Czamara, D., Pehl, V., Wagenbuechler, P., Feldmann, L., Quickenstedt-Reinhardt, P., Allgaier, A.-K., Freisleder, F. J., Greimel, E., Kvist, T., Lahti, J., Räikkönen, K., Rex-Haffner, M., Arnarson, E.Ö., Craighead, W. E., Schulte-Körne, G., & Binder, E. B. (2019). Polygenic Risk: Predicting Depression Outcomes in Clinical and Epidemiological Cohorts of Youths. American Journal of Psychiatry, 176(8), 615–625. https://doi.org/10.1176/appi.ajp.2019.18091014

Howard, D. M., Adams, M. J., Clarke, T. K., Hafferty, J. D., Gibson, J., Shirali, M., Coleman, J. R. I., Hagenaars, S. P., Ward, J., Wigmore, E. M., Alloza, C., Shen, X., Barbu, M. C., Xu, E. Y., Whalley, H. C., Marioni, R. E., Porteous, D. J., Davies, G., Deary, I. J., … McIntosh, A. M. (2019). Genome-wide meta-analysis of depression identifies 102 independent variants and highlights the importance of the prefrontal brain regions. Nature Neuroscience, 22(3), 343–352. https://doi.org/10.1038/s41593-018-0326-7

Jansen, R., Batista, S., Brooks, A. I., Tischfield, J. A., Willemsen, G., van Grootheest, G., Hottenga, J.-J., Milaneschi, Y., Mbarek, H., Madar, V., Peyrot, W., Vink, J. M., Verweij, C. L., de Geus, E. J., Smit, J. H., Wright, F. A., Sullivan, P. F., Boomsma, D. I., & Penninx, B. W. (2014). Sex differences in the human peripheral blood transcriptome. BMC Genomics, 15(1), 33. https://doi.org/10.1186/1471-2164-15-33

Jessen, H. M., & Auger, A. P. (2011). Sex differences in epigenetic mechanisms may underlie risk and resilience for mental health disorders. Epigenetics, 6(7), 857–861. http://www.ncbi.nlm.nih.gov/pubmed/21617370

Johnson, W. E., Li, C., & Rabinovic, A. (2007). Adjusting batch effects in microarray expression data using empirical Bayes methods. Biostatistics (Oxford, England), 8(1), 118–127. https://doi.org/10.1093/biostatistics/kxj037

Kang, H. J., Park, Y., Yoo, K. H., Kim, K. T., Kim, E. S., Kim, J. W., Kim, S. W., Shin, I. S., Yoon, J. S., Kim, J. H., & Kim, J. M. (2020). Sex differences in the genetic architecture of depression. Scientific Reports, 10(1), 1–12. https://doi.org/10.1038/s41598-020-66672-9

Karisetty, B. C., Khandelwal, N., Kumar, A., & Chakravarty, S. (2017). Sex difference in mouse hypothalamic transcriptome profile in stress-induced depression model. Biochemical and Biophysical Research Communications, 486(4), 1122–1128. https://doi.org/10.1016/j.bbrc.2017.04.005

Kelly, M. M., Tyrka, A. R., Anderson, G. M., Price, L. H., & Carpenter, L. L. (2008). Sex differences in emotional and physiological responses to the Trier Social Stress Test. Journal of Behavior Therapy and Experimental Psychiatry, 39(1), 87–98. https://doi.org/10.1016/j.jbtep.2007.02.003

Khramtsova, E. A., Davis, L. K., & Stranger, B. E. (2019). The role of sex in the genomics of human complex traits. Nature Reviews Genetics, 20(3), 173–190. https://doi.org/10.1038/s41576-018-0083-1

Labonté, B., Engmann, O., Purushothaman, I., Menard, C., Wang, J., Tan, C., Scarpa, J. R., Moy, G., Loh, Y.-H. E., Cahill, M., Lorsch, Z. S., Hamilton, P. J., Calipari, E. S., Hodes, G. E., Issler, O., Kronman, H., Pfau, M., Obradovic, A. L. J., Dong, Y., … Nestler, E. J. (2017). Sex-specific transcriptional signatures in human depression. Nature Medicine, 23(9), 1102–1111. https://doi.org/10.1038/nm.4386

Lee, J. J., Wedow, R., Okbay, A., Kong, E., Maghzian, O., Zacher, M., Nguyen-Viet, T. A., Bowers, P., Sidorenko, J., Karlsson Linnér, R., Fontana, M. A., Kundu, T., Lee, C., Li, H., Li, R., Royer, R., Timshel, P. N., Walters, R. K., Willoughby, E. A., … Turley, P. (2018). Gene discovery and polygenic prediction from a genome-wide association study of educational attainment in 1.1 million individuals. Nature Genetics, 50(8), 1112–1121. https://doi.org/10.1038/s41588-018-0147-3

Lee, P. H., Anttila, V., Won, H., Feng, Y. C. A., Rosenthal, J., Zhu, Z., Tucker-Drob, E. M., Nivard, M. G., Grotzinger, A. D., Posthuma, D., Wang, M. M. J., Yu, D., Stahl, E. A., Walters, R. K., Anney, R. J. L., Duncan, L. E., Ge, T., Adolfsson, R., Banaschewski, T., … Smoller, J. W. (2019). Genomic Relationships, Novel Loci, and Pleiotropic Mechanisms across Eight Psychiatric Disorders. Cell, 179(7), 1469-1482.e11. https://doi.org/10.1016/j.cell.2019.11.020

Leek, J. T., Johnson, W. E., Parker, H. S., Jaffe, A. E., & Storey, J. D. (2012). The sva package for removing batch effects and other unwanted variation in high-throughput experiments. Bioinformatics, 28(6), 882–883. https://doi.org/10.1093/bioinformatics/bts034

Lindén, M., Ramírez Sepúlveda, J. I., James, T., Thorlacius, G. E., Brauner, S., Gómez-Cabrero, D., Olsson, T., Kockum, I., & Wahren-Herlenius, M. (2017). Sex influences eQTL effects of SLE and Sjögren’s syndrome-associated genetic polymorphisms. Biology of Sex Differences, 8(1), 34. https://doi.org/10.1186/s13293-017-0153-7

Liu, J. J. W., Ein, N., Peck, K., Huang, V., Pruessner, J. C., & Vickers, K. (2017). Sex differences in salivary cortisol reactivity to the Trier Social Stress Test (TSST): A meta-analysis. Psychoneuroendocrinology, 82, 26–37. https://doi.org/10.1016/j.psyneuen.2017.04.007

Matthews, S. G. (1998). Dynamic changes in glucocorticoid and mineralocorticoid receptor mRNA in the developing guinea pig brain. Developmental Brain Research, 107(1), 123–132. https://doi.org/10.1016/S0165-3806(98)00008-X

Mayberg, H. S., Lozano, A. M., Voon, V., McNeely, H. E., Seminowicz, D., Hamani, C., Schwalb, J. M., & Kennedy, S. H. (2005). Deep brain stimulation for treatment-resistant depression. Neuron, 45(5), 651–660. https://doi.org/10.1016/j.neuron.2005.02.014

Mayne, B. T., Bianco-Miotto, T., Buckberry, S., Breen, J., Clifton, V., Shoubridge, C., & Roberts, C. T. (2016). Large Scale Gene Expression Meta-Analysis Reveals Tissue-Specific, Sex-Biased Gene Expression in Humans. Frontiers in Genetics, 7, 183. https://doi.org/10.3389/fgene.2016.00183

Moore, S. R. (2017). Commentary: What is the case for candidate gene approaches in the era of high-throughput genomics? A response to Border and Keller (2017). Journal of Child Psychology and Psychiatry, 58(3), 331–334. https://doi.org/10.1111/jcpp.12697

Morrison, K. E., Rodgers, A. B., Morgan, C. P., & Bale, T. L. (2014). Epigenetic mechanisms in pubertal brain maturation. Neuroscience, 264, 17–24. https://doi.org/10.1016/j.neuroscience.2013.11.014

Muglia, P., Tozzi, F., Galwey, N. W., Francks, C., Upmanyu, R., Kong, X. Q., Antoniades, A., Domenici, E., Perry, J., Rothen, S., Vandeleur, C. L., Mooser, V., Waeber, G., Vollenweider, P., Preisig, M., Lucae, S., Müller-Myhsok, B., Holsboer, F., Middleton, L. T., & Roses, A. D. (2010). Genome-wide association study of recurrent major depressive disorder in two European case-control cohorts. Molecular Psychiatry, 15(6), 589–601. https://doi.org/10.1038/mp.2008.131

Nievergelt, C. M., Maihofer, A. X., Klengel, T., Atkinson, E. G., Chen, C. Y., Choi, K. W., Coleman, J. R. I., Dalvie, S., Duncan, L. E., Gelernter, J., Levey, D. F., Logue, M. W., Polimanti, R., Provost, A. C., Ratanatharathorn, A., Stein, M. B., Torres, K., Aiello, A. E., Almli, L. M., … Koenen, K. C. (2019). International meta-analysis of PTSD genome-wide association studies identifies sex- and ancestry-specific genetic risk loci. Nature Communications, 10(1). https://doi.org/10.1038/s41467-019-12576-w

Owen, D., & Matthews, S. G. (2003). Glucocorticoids and sex-dependent development of brain glucocorticoid and mineralocorticoid receptors. Endocrinology, 144(7), 2775– 2784. https://doi.org/10.1210/en.2002-0145

Pasman, J. A., Verweij, K. J. H., Gerring, Z., Stringer, S., Sanchez-Roige, S., Treur, J. L., Abdellaoui, A., Nivard, M. G., Baselmans, B. M. L., Ong, J. S., Ip, H. F., van der Zee, M. D., Bartels, M., Day, F. R., Fontanillas, P., Elson, S. L., de Wit, H., Davis, L. K., MacKillop, J., … Vink, J. M. (2018). GWAS of lifetime cannabis use reveals new risk loci, genetic overlap with psychiatric traits, and a causal influence of schizophrenia. Nature Neuroscience, 21(9), 1161–1170. https://doi.org/10.1038/s41593-018-0206-1

Pruitt, K. D., Tatusova, T., Brown, G. R., & Maglott, D. R. (2012). NCBI Reference Sequences (RefSeq): current status, new features and genome annotation policy. Nucleic Acids Research, 40(Database issue), D130–5. https://doi.org/10.1093/nar/gkr1079

Ramikie, T. S., & Ressler, K. J. (2018). Mechanisms of Sex Differences in Fear and Posttraumatic Stress Disorder. Biological Psychiatry, 83(10), 876–885. https://doi.org/10.1016/J.BIOPSYCH.2017.11.016

Reul, J. M. H. M., & De Kloet, E. R. (1985). Two receptor systems for corticosterone in rat brain: Microdistribution and differential occupation. Endocrinology, 117(6), 2505–2511. https://doi.org/10.1210/endo-117-6-2505

Ripke, S., Neale, B. M., Corvin, A., Walters, J. T. R., Farh, K. H., Holmans, P. A., Lee, P., Bulik-Sullivan, B., Collier, D. A., Huang, H., Pers, T. H., Agartz, I., Agerbo, E., Albus, M., Alexander, M., Amin, F., Bacanu, S. A., Begemann, M., Belliveau, R. A., … O’Donovan, M. C. (2014). Biological insights from 108 schizophrenia-associated genetic loci. Nature, 511(7510), 421–427. https://doi.org/10.1038/nature13595

Ripke, S., Wray, N. R., Lewis, C. M., Hamilton, S. P., Weissman, M. M., Breen, G., Byrne, E. M., Blackwood, D. H. R., Boomsma, D. I., Cichon, S., Heath, A. C., Holsboer, F., Lucae, S., Madden, P. A. F., Martin, N. G., McGuffin, P., Muglia, P., Noethen, M. M., Penninx, B. P., … Sullivan, P. F. (2013). A mega-analysis of genome-wide association studies for major depressive disorder. Molecular Psychiatry, 18(4), 497–511. https://doi.org/10.1038/mp.2012.21

Rowson, S. A., Bekhbat, M., Kelly, S. D., Binder, E. B., Hyer, M. M., Shaw, G., Bent, M. A., Hodes, G., Tharp, G., Weinshenker, D., Qin, Z., & Neigh, G. N. (2019). Chronic adolescent stress sex-specifically alters the hippocampal transcriptome in adulthood. Neuropsychopharmacology, 44(7), 1207–1215. https://doi.org/10.1038/s41386-019-0321-z

Salk, R. H., Hyde, J. S., & Abramson, L. Y. (2017). Gender differences in depression in representative national samples: Meta-analyses of diagnoses and symptoms. Psychological Bulletin, 143(8), 783–822. https://doi.org/10.1037/bul0000102

Santarelli, S., Zimmermann, C., Kalideris, G., Lesuis, S. L., Arloth, J., Uribe, A., Dournes, C., Balsevich, G., Hartmann, J., Masana, M., Binder, E. B., Spengler, D., & Schmidt, M. V. (2017). An adverse early life environment can enhance stress resilience in adulthood. Psychoneuroendocrinology, 78, 213–221. https://doi.org/10.1016/j.psyneuen.2017.01.021

Sapolsky, R. M., Romero, L. M., & Munck, A. U. (2000). How Do Glucocorticoids Influence Stress Responses? Integrating Permissive, Suppressive, Stimulatory, and Preparative Actions ^1^. Endocrine Reviews, 21(1), 55–89. https://doi.org/10.1210/edrv.21.1.0389

Savage, J. E., Jansen, P. R., Stringer, S., Watanabe, K., Bryois, J., De Leeuw, C. A., Nagel, M., Awasthi, S., Barr, P. B., Coleman, J. R. I., Grasby, K. L., Hammerschlag, A. R., Kaminski, J. A., Karlsson, R., Krapohl, E., Lam, M., Nygaard, M., Reynolds, C. A., Trampush, J. W., … Posthuma, D. (2018). Genome-wide association meta-analysis in 269,867 individuals identifies new genetic and functional links to intelligence. Nature Genetics, 50(7), 912–919. https://doi.org/10.1038/s41588-018-0152-6

Shabalin, A. A. (2012). Matrix eQTL: Ultra fast eQTL analysis via large matrix operations. Bioinformatics, 28(10), 1353–1358. https://doi.org/10.1093/bioinformatics/bts163

Smoller, J. W., Kendler, K., Craddock, N., Lee, P. H., Neale, B. M., Nurnberger, J. N., Ripke, S., Santangelo, S., Sullivan, P. S., Neale, B. N., Purcell, S., Anney, R., Buitelaar, J., Fanous, A., Faraone, S. F., Hoogendijk, W., Lesch, K. P., Levinson, D. L., Perlis, R. P., … O’Donovan, M. (2013). Identification of risk loci with shared effects on five major psychiatric disorders: A genome-wide analysis. The Lancet, 381(9875), 1371–1379. https://doi.org/10.1016/S0140-6736(12)62129-1

Stahl, E. A., Breen, G., Forstner, A. J., McQuillin, A., Ripke, S., Trubetskoy, V., Mattheisen, M., Wang, Y., Coleman, J. R. I., Gaspar, H. A., de Leeuw, C. A., Steinberg, S., Pavlides, J. M. W., Trzaskowski, M., Byrne, E. M., Pers, T. H., Holmans, P. A., Richards, A. L., Abbott, L., … Sklar, P. (2019). Genome-wide association study identifies 30 loci associated with bipolar disorder. Nature Genetics, 51(5), 793–803. https://doi.org/10.1038/s41588-019-0397-8

Stephens, M. A. C., Mahon, P. B., McCaul, M. E., & Wand, G. S. (2016). Hypothalamic– pituitary–adrenal axis response to acute psychosocial stress: Effects of biological sex and circulating sex hormones. Psychoneuroendocrinology, 66, 47–55. https://doi.org/10.1016/J.PSYNEUEN.2015.12.021

Sugathan, A., & Waxman, D. J. (2013). Genome-wide analysis of chromatin states reveals distinct mechanisms of sex-dependent gene regulation in male and female mouse liver. Molecular and Cellular Biology, 33(18), 3594–3610. https://doi.org/10.1128/MCB.00280-13

Terada, A., Tsuda, K., & Sese, J. (2013). Fast Westfall-Young permutation procedure for combinatorial regulation discovery. Proceedings - 2013 IEEE International Conference on Bioinformatics and Biomedicine, IEEE BIBM 2013, 153–158. https://doi.org/10.1109/BIBM.2013.6732479

Tiwari, A., & Gonzalez, A. (2018). Biological alterations affecting risk of adult psychopathology following childhood trauma: A review of sex differences. In Clinical Psychology Review (Vol. 66, pp. 69–79). Elsevier Inc. https://doi.org/10.1016/j.cpr.2018.01.006

Westra, H. J., Peters, M. J., Esko, T., Yaghootkar, H., Schurmann, C., Kettunen, J., Christiansen, M. W., Fairfax, B. P., Schramm, K., Powell, J. E., Zhernakova, A., Zhernakova, D. V., Veldink, J. H., Van Den Berg, L. H., Karjalainen, J., Withoff, S., Uitterlinden, A. G., Hofman, A., Rivadeneira, F., … Franke, L. (2013). Systematic identification of trans eQTLs as putative drivers of known disease associations. Nature Genetics, 45(10), 1238–1243. https://doi.org/10.1038/ng.2756

Wray, N. R., Ripke, S., Mattheisen, M., Trzaskowski, M., Byrne, E. M., Abdellaoui, A., Adams, M. J., Agerbo, E., Air, T. M., Andlauer, T. M. F., Bacanu, S.-A., Bækvad-Hansen, M., Beekman, A. F. T., Bigdeli, T. B., Binder, E. B., Blackwood, D. R. H., Bryois, J., Buttenschøn, H. N., Bybjerg-Grauholm, J., … Sullivan, P. F. (2018a). Genome-wide association analyses identify 44 risk variants and refine the genetic architecture of major depression. Nature Genetics, 50(5), 668–681. https://doi.org/10.1038/s41588-018-0090-3

Wray, N. R., Ripke, S., Mattheisen, M., Trzaskowski, M., Byrne, E. M., Abdellaoui, A., Adams, M. J., Agerbo, E., Air, T. M., Andlauer, T. M. F., Bacanu, S.-A., Bækvad-Hansen, M., Beekman, A. F. T., Bigdeli, T. B., Binder, E. B., Blackwood, D. R. H., Bryois, J., Buttenschøn, H. N., Bybjerg-Grauholm, J., … Sullivan, P. F. (2018b). Genome-wide association analyses identify 44 risk variants and refine the genetic architecture of major depression. Nature Genetics, 50(5), 668–681. https://doi.org/10.1038/s41588-018-0090-3

Xue, A., Wu, Y., Zhu, Z., Zhang, F., Kemper, K. E., Zheng, Z., Yengo, L., Lloyd-Jones, L. R., Sidorenko, J., Wu, Y., Agbessi, M., Ahsan, H., Alves, I., Andiappan, A., Awadalla, P., Battle, A., Beutner, F., Bonder, M. J. J., Boomsma, D., … Yang, J. (2018). Genome-wide association analyses identify 143 risk variants and putative regulatory mechanisms for type 2 diabetes. Nature Communications, 9(1), 1–14. https://doi.org/10.1038/s41467-018-04951-w

Yao, C., Joehanes, R., Johnson, A. D., Huan, T., Esko, T., Ying, S., Freedman, J. E., Murabito, J., Lunetta, K. L., Metspalu, A., Munson, P. J., & Levy, D. (2014). Sex- and age-interacting eQTLs in human complex diseases. Human Molecular Genetics, 23(7), 1947–1956. https://doi.org/10.1093/hmg/ddt582

Yu, D., Sul, J. H., Tsetsos, F., Nawaz, M. S., Huang, A. Y., Zelaya, I., Illmann, C., Osiecki, L., Darrow, S. M., Hirschtritt, M. E., Greenberg, E., Muller-Vahl, K. R., Stuhrmann, M., Dion, Y., Rouleau, G. A., Aschauer, H., Stamenkovic, M., Schlögelhofer, M., Sandor, P., … Worbe, Y. (2019). Interrogating the genetic determinants of Tourette’s syndrome and other tiC disorders through genome-wide association studies. American Journal of Psychiatry, 176(3), 217–227. https://doi.org/10.1176/appi.ajp.2018.18070857

Zhou, J., & Troyanskaya, O. G. (2015). Predicting effects of noncoding variants with deep learning-based sequence model. Nature Methods, 12(10), 931–934. https://doi.org/10.1038/nmeth.3547

Zimmermann, C. A., Arloth, J., Santarelli, S., Löschner, A., Weber, P., Schmidt, M. V., Spengler, D., & Binder, E. B. (2019). Stress dynamically regulates co-expression networks of glucocorticoid receptor-dependent MDD and SCZ risk genes. Translational Psychiatry, 9(1), 41. https://doi.org/10.1038/s41398-019-0373-1

Zorn, J. V., Schür, R. R., Boks, M. P., Kahn, R. S., Joëls, M., & Vinkers, C. H. (2017). Cortisol stress reactivity across psychiatric disorders: A systematic review and meta-analysis. Psychoneuroendocrinology, 77, 25–36. https://doi.org/10.1016/J.PSYNEUEN.2016.11.036

## References

Arloth, J., Bogdan, R., Weber, P., Frishman, G., Menke, A., Wagner, K. V., Balsevich, G., Schmidt, M. V., Karbalai, N., Czamara, D., Altmann, A., Trümbach, D., Wurst, W., Mehta, D., Uhr, M., Klengel, T., Erhardt, A., Carey, C. E., Conley, E. D., … Binder, E.B. (2015). Genetic Differences in the Immediate Transcriptome Response to Stress Predict Risk-Related Brain Function and Psychiatric Disorders. Neuron, 86(5), 1189– 1202. https://doi.org/10.1016/j.neuron.2015.05.034

Consortium, T. Gte. (2020). The GTEx Consortium atlas of genetic regulatory effects across human tissues. Science, 369(6509), 1318–1330. https://doi.org/10.1126/science.aaz1776

Zhernakova, D. V., Deelen, P., Vermaat, M., Van Iterson, M., Van Galen, M., Arindrarto, W., Van’t Hof, P., Mei, H., Van Dijk, F., Westra, H. J., Bonder, M. J., Van Rooij, J., Verkerk, M., Jhamai, P. M., Moed, M., Kielbasa, S. M., Bot, J., Nooren, I., Pool, R., … Franke, L. (2017). Identification of context-dependent expression quantitative trait loci in whole blood. Nature Genetics, 49(1), 139–145. https://doi.org/10.1038/ng.3737

